# Lamina Cribrosa Strain as a Glaucoma Prognostic Factor: The STRAIN Study Methodology and Initial Outcomes

**DOI:** 10.64898/2026.11.09.26367640

**Authors:** Patricia M. Sabin, Farzaneh Dadvar, Rebecca Mirville, Yulia Nam, Fatma Hamid, Chris Bradley, Jithin Yohannan, Kelly A. Clingo, Thomas V. Johnson, Bryce Chiang, Harry A. Quigley, Thao D. Nguyen

## Abstract

**Objective:** To estimate the lamina cribrosa (LC) strain response to intraocular pressure (IOP) change in open angle glaucoma (OAG) patients and examine associations between LC strain and demographic and clinical characteristics relevant to OAG in an ongoing longitudinal study.

**Design:** Cohort study

**Participants:** Adult patients with OAG (205 eyes of 119 patients)

**Methods:** The STRAIN study is an ongoing longitudinal study designed to determine whether LC biomechanical strain is associated with present, prior, and future OAG damage. In this initial analysis, patients either temporarily stopped ≥1 eye drop, initiated an eye drop, or underwent selective laser trabeculoplasty to produce IOP change. Optical coherence tomography (OCT) scans of the optic nerve head (ONH) acquired before and up to several weeks after IOP change were analyzed using digital volume correlation (DVC) to estimate LC strain. Strain measurements were compared with age, race, sex, axial length, central corneal thickness (CCT), corneal hysteresis, optic disc area, years of known OAG, retinal nerve fiber layer (RNFL) thickness, and visual field mean deviation (MD).

**Main Outcome Measures:** LC strain estimated by DVC from OCT scans acquired before and after IOP alteration.

**Results:** Axial LC strain (*Ezz*) was tensile with IOP decrease and compressive with IOP increase. Maximum principal strain was tensile (mean *E_max_* = 1.09%), and maximum shear strain was positive (mean *Γ_max_* = 1.16%) in the radial-axial plane. Both increased quadratically with the IOP change. Older age was significantly related to greater tensile circumferential strain (*p* = 0.02) and lower *Γ_max_* (*p* = 0.01). Sex, CCT, corneal hysteresis, and optic disc area were not significantly associated with LC strain. In combined multivariable models adjusting for IOP change, demographics, and accounting for correlations between eyes from the same patient, eyes with thinner inferior-quadrant RNFL had smaller *Γ_max_* for a given IOP change (*p* = 0.02).

**Conclusions:** LC strain in patients with OAG is associated with the degree of neuronal loss, age, and axial length, supporting a relationship between the LC biomechanical response and factors relevant to glaucomatous damage.

## 1. Introduction

Open angle glaucoma (OAG) is a complex and heterogeneous disease with multiple phenotypes.^1,2^ Prospective studies have identified several prognostic factors^3^ associated with the onset and progression of OAG, including older age, higher intraocular pressure (IOP), African derivation, and family history.^4–6^ Additional factors, including myopia and large optic disc area, have been associated with OAG in cross-sectional studies, although their prognostic significance is less established. However, the factors associated with OAG risk and progression may differ among disease phenotypes, including those characterized by different IOP levels at onset.^7^ Such phenotype-dependent differences may reflect variation in the biomechanical response of the eye to IOP. Identifying additional factors that characterize the susceptibility of an individual to glaucomatous damage may therefore improve understanding of disease mechanisms and ultimately provide clinically useful prognostic factors.

The biomechanical behavior of the lamina cribrosa (LC) may be important to glaucoma susceptibility. The LC is a connective tissue structure within the optic nerve head (ONH) that provides structural and physiological support for retinal ganglion cell (RGC) axons as they exit the eye. Studies of human OAG eyes and animal models of ocular hypertension have demonstrated injury to RGC axons at the LC,^8^ and IOP-induced stress and deformation of the LC may contribute to this injury. Tissue deformation and remodeling of the ONH are established features of glaucomatous optic neuropathy,^9–12^ and biomechanical models have suggested that differences in stress and strain within the LC may be related to glaucoma susceptibility.^13,14^ Therefore, the mechanical response of the LC to changes in IOP may provide information about the biomechanical susceptibility of individual eyes to glaucomatous damage.

Studies of *postmortem* human eyes have provided important evidence regarding the biomechanical behavior of the LC. Inflation testing has demonstrated regional variation in pressure-induced LC strain across anatomical quadrants and between central and peripheral regions.^15–18^ LC strain has also been shown to vary with age, with lower strains observed in older eyes.^17^ Studies comparing eyes with and without glaucoma have found differences in the LC pressure-strain response,^18,19^ and the relationship between LC strain and the extent of optic nerve damage may be nonmonotonic.^19^ In addition, differences in the LC biomechanical response have been observed among racial groups.^20^ However, *postmortem* studies cannot capture the mechanical behavior of the living ONH, where tissue hydration, composition, blood flow, and remodeling may differ. Measurement of LC biomechanics in living eyes is therefore important for determining whether responses observed *ex vivo* are also present in patients with glaucoma.

Advances in optical coherence tomography (OCT) imaging have enabled noninvasive measurement of ONH deformation and strain in living human eyes. Several studies have used OCT-based imaging, including digital volume correlation (DVC) and related approaches, to characterize LC deformation following clinically induced IOP changes.^21–30^ Our previous studies measured the *in vivo* LC strain response following IOP elevation using tight-fitting goggles and IOP reduction following laser suture lysis of trabeculectomy flaps or glaucoma medication changes, demonstrating that the strain response varies with the magnitude and duration of IOP change.^21–24^ In contrast, Girard et al. found no significant association between the magnitude of IOP reduction and LC strain response following trabeculectomy.^25^ Studies have also demonstrated relationships between LC or ONH strain and glaucomatous damage. Our previous studies found greater LC strain responses with worse visual field indices and thinner RNFL.^22,24^ Girard et al. found greater LC response following trabeculectomy in eyes with worse visual field damage.^25^ More recently, Chuangsuwanich et al. found an association between higher ONH strain induced by ophthalmodynamometric IOP elevation and lower retinal sensitivity in glaucoma patients with higher baseline IOP than those with normal baseline IOP.^26^ Differences in the methods of IOP alteration, imaging protocols, and duration of IOP change may contribute to variability among these findings. IOP elevation caused by ophthalmodynamometer indentation may push the globe into the orbital tissues and produce a different strain field in the ONH than IOP elevation caused by altering aqueous humor dynamics. Similar to Girard et al.,^25^ our prior study applying tight-fitting goggles to push on the eye and elevate IOP also found no significant association between LC strain and IOP change.^23^ A clinical design that measures the LC strain response to both increasing and decreasing IOP by altering aqueous humor dynamics may help clarify these relationships.

The Strain Assessment In glaucomatous Neuropathy (STRAIN) study is a prospective clinical trial designed to investigate LC strain as a potential prognostic biomarker in OAG. For the baseline assessment, we used spectral domain OCT to image the ONH region in a large cohort of OAG patients before and several weeks after an intervention to alter IOP, either through starting or stopping a glaucoma medication or selective laser trabeculoplasty (SLT). Digital volume correlation (DVC) was used to analyze the pre- and post-treatment OCT images and estimate the LC strain response to IOP change. Here, we report strain measurements and examine their cross-sectional associations with demographic and ocular parameters, including the degree of OAG damage assessed by retinal nerve fiber layer (RNFL) thickness and visual field measurements. Our earlier studies, Czerpak et al.^22,23^ and Hannay et al.^24^, also investigated cross-sectional associations between LC strains and OAG damage; however, those were limited by a significantly smaller sample size disproportionately represented by moderate to severe disease. In future analyses, we will compare these strain measurements with measures of retrospective damage progression from existing electronic data and with prospective OCT and visual field measurements collected over three years of follow-up.

## 2. Methods

### 2.1 Study approval

This research was registered as a clinical trial at ClinicalTrials.gov (NCT06240312: Optic Nerve Head Strain as Biomarker for Glaucoma, registered 06-11-2025, sponsor: Johns Hopkins University). It was approved and is monitored by the Institutional Review Board of the Johns Hopkins School of Medicine, and participants signed written consent to participate. The study abides by the Declaration of Helsinki. Participants were active patients of several glaucoma specialists at the Glaucoma Center of Excellence at the Wilmer Eye Institute, with recruitment starting in August 2024. Eligible candidates were informed of the study during regular clinic visits by their glaucoma specialist and referred to the research coordinator.

### 2.2 Participant selection

Eligible participants were >30 and <90 years of age from any race (derivation) with OAG defined as satisfying, in at least one eye, the Iyer et al. criteria,^1^ namely: 1) an RNFL OCT quadrant that is statistically outside the normal range, 2) a matching HFA3 glaucoma hemifield test (GHT) outside normal limits with its defect in the opposite hemifield from the OCT quadrant defect, and 3) a clinical retinal and ONH examination that exhibited no non-glaucomatous finding that could explain the OCT and field findings. Both eyes were included if at least one eye met inclusion criteria. Gonioscopy by a glaucoma specialist assured that the angle was open and free of secondary glaucoma features.

Subjects were excluded if they had physical inability to sit upright for imaging, had significant media opacity or ocular condition impairing imaging, could not tolerate imaging or tonometry, had unreliable OCT or field testing, had a secondary form of glaucoma, had myopia >8 diopters, had field mean deviation (MD) worse than −12 dB in the better eye, were recognized to need immediate glaucoma surgery, had eye surgery within 3 months of baseline exams, had an ocular condition that could affect either RNFL or field (such as age-related macular degeneration, retinal vascular occlusion, or diabetic retinopathy), had a history of corneal disease that could compromise tonometry or imaging, or had a language or hearing impairment affecting communication or understanding of the research protocol.

### 2.3 IOP change methods

To examine the LC strain response to IOP change in both directions, study participants were assigned to one of two groups. The **washout group** consisted of patients taking one or more topical glaucoma eye drops at the time of enrollment. To increase IOP, these subjects were asked to temporarily discontinue one or more of these medications. Before discontinuing medication, patients measured their IOP at home for one week while continuing their usual medications, using the iCare HOME2 device as detailed below. They then discontinued one or more eye drops for a period sufficient to establish that their IOP had increased, which varied from one week to more than one month (Figure 1). For patients taking multiple types of eye drops at baseline, prostaglandin was discontinued first. If IOP did not increase during monitoring after discontinuation of the first medication, additional medications were discontinued sequentially until an IOP increase was observed or all medications had been stopped. In a small number of eyes (*n* = 14), IOP did not increase even after discontinuation of all drops. After the washout period, subjects returned for a second imaging session at the higher IOP and subsequently restarted the discontinued medication(s). The median time between imaging for washout patients was 32 days. The data from home tonometry are being prepared for a separate publication.

**Figure 1:**
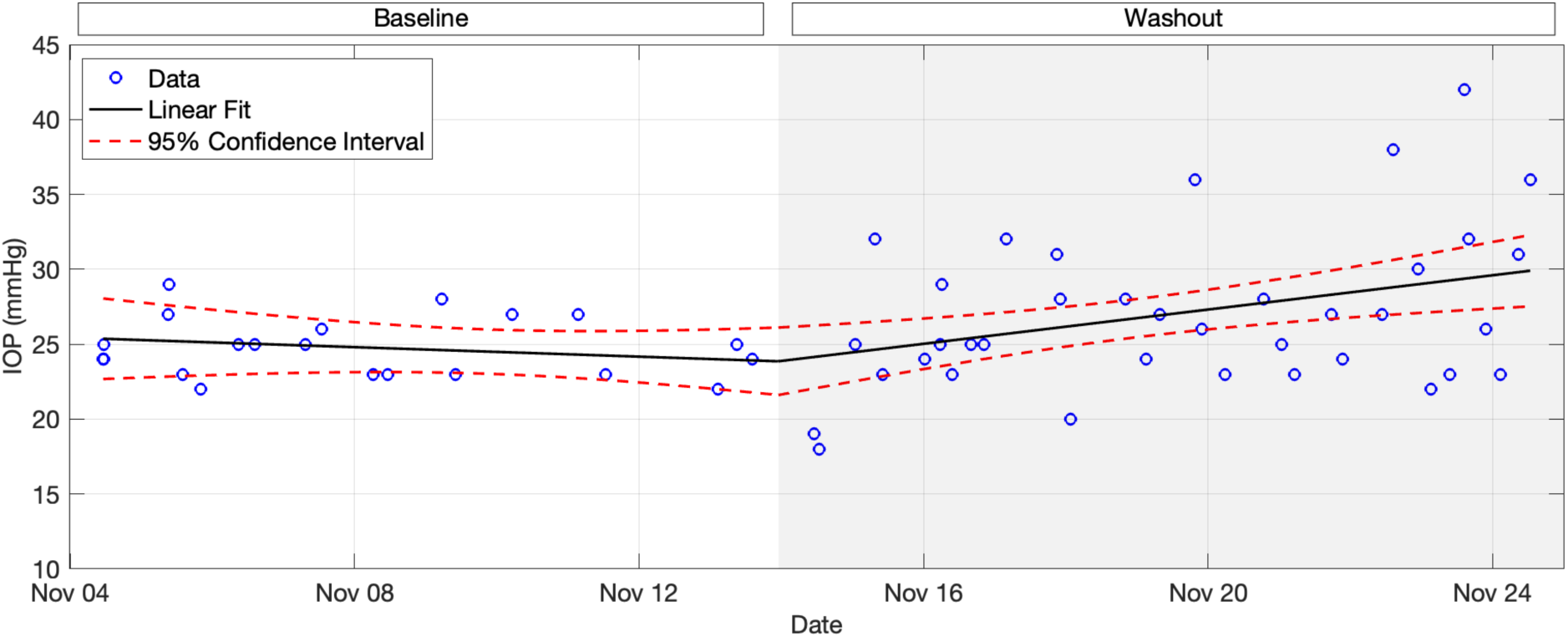
Example IOP measurements using home tonometry. IOP measurements from one representative patient are shown. Patients measured their IOP at home using an iCare HOME2 device. Baseline IOP was established while continuing their usual medications for one week. Following baseline monitoring, one or more eye drops were discontinued and home IOP monitoring continued until an increase in IOP was established.

The **wash-in group** underwent IOP lowering, either through initiation of a glaucoma eye drop or treatment with SLT. The study protocol did not prescribe a treatment; rather, the treating physician and patient selected the treatment plan as part of their routine glaucoma care. In most cases, the eye drop initiated was a prostaglandin (Supplemental Table S1). Approximately half of the subjects in the wash-in group also monitored their IOP with home tonometry while initiating the new eye drop.

### 2.4 Testing procedures

With the exception of the period of eye drop discontinuation in the washout group, all clinical care and treatment decisions were made by the physician and patient as part of routine glaucoma management. This included outpatient visits every 3-6 months under typical care, or more frequently as indicated by the course of disease. At each follow-up visit, changes in treatment, including medication adjustments or surgical interventions, were recorded. All changes in therapy occurring during the planned 3-year follow-up will be incorporated into the final analysis following completion of prospective data collection.

#### 2.4.1 Keratometry and axial length measurements

Keratometry and axial length measurements were acquired using either the IOLMaster (Zeiss Meditec, Dublin, CA, USA) or the Refracto-keratometer (LIINNCXM, Shanghai Link Instruments, Ltd., China). Keratometry values were entered into the OCT system prior to each imaging session to derive the appropriate optical magnification and scaling factors. Since a proportion of study patients were pseudophakic, the original refractive error could not be determined, and we used axial length as a surrogate.

#### 2.4.2 OCT imaging

OCT images were acquired using the Spectralis OCT system with HEYEX software (Heidelberg Engineering GmbH, Heidelberg, Germany), as previously described.^21^ Briefly, the radial 24-scan protocol, centered on the optic disc, was used to capture the ONH volume. Radial scans had a resolution of 768 × 495 pixels in the (R,Z) plane, with each image generated from an average of 25 B-scans and 768 A-scans per B-scan in enhanced depth imaging mode. Resolution in the anterior-posterior (Z) direction was 3.87 μm/pixel, while resolution in the radial (R) direction varied from 5.18 to 6.98 μm/pixel. Patients were imaged three times at each session, with 30 seconds between acquisitions. We exported detailed image data using software provided by Heidelberg. For RNFL thickness analysis, the global and 4-quadrant thickness measurements and their statistical comparison with the normative database were extracted from each image session.

#### 2.4.3 IOP measurements

At each imaging session, a Glaucoma Center certified ophthalmic technician or physician measured IOP by Goldmann applanation tonometry as part of the standard clinic visit, and the research coordinator obtained six consecutive IOP measurements using an iCare HOME2 tonometer (TA023; iCare Finland Oy, Vantaa, Finland) to generate a mean in-office iCare IOP value. iCare measurements were accepted when there was good or excellent agreement among the six measurements. The instrument automatically excludes the lowest and highest measurements for added reliability.

For home tonometry, patients used the iCare HOME2 tonometer. All subjects in the washout group were offered training and access to a home tonometer. Subjects in the wash-in group who were initiating a new eye drop and expressed interest in home tonometry were also offered training. Wash-in group subjects who underwent selective laser trabeculoplasty did not perform home tonometry and were examined 6 weeks after treatment.

Patients performing self-tonometry received in-office training from a research coordinator and were required to demonstrate accurate self-measurement, defined as obtaining three consecutive self-measurements within 5 mm Hg of a concomitant Goldmann applanation measurement and within a range of <7 mm Hg across the three self-measurements, before taking the device home. Patients were then instructed to obtain 2-4 daytime measurements per day during the testing period. Real-time home iCare measurements were available to both the patient and the research coordinator, and the research coordinator monitored these measurements to assess whether a sufficient IOP change had occurred and to ensure that IOP remained within a safe range. A mean IOP change of at least 3 mm Hg was targeted before scheduling post-intervention imaging. This threshold was based on previous work demonstrating that DVC analysis of OCT volumes could detect mean LC axial strain that was statistically greater than the mean baseline strain error for IOP changes >2.8 mm Hg.^21^

#### 2.4.4 Central corneal thickness and hysteresis

In each eye, CCT was measured at the pre-intervention visit using an Ultrasonic Pachymeter (DGH Technology, Inc., Exton, PA, USA). Corneal hysteresis was measured at both the pre- and post-intervention visits using an Ocular Response Analyzer (Reichert, Inc, Depew, NY).

#### 2.4.5 Visual field data

Visual field testing was performed using Zeiss HFA3 instruments with the SITA-Faster algorithm (Carl Zeiss Meditec, Dublin, CA). Three visual fields were obtained for each eye during the baseline period and at least every 6 months thereafter.

Additional sets of three visual fields per eye are planned at 2 and at 3 years after baseline, all using the SITA-Faster algorithm.

#### 2.4.6 Retrospective data analysis

Most patients had undergone several years of prior care at the Glaucoma Center of Excellence. The study approval permitted review and analysis of subjects’ electronic medical records for the following features: age, sex, self-described race/derivation/ethnicity, duration of known OAG diagnosis, past and present glaucoma medications and procedural treatments, IOP measurements recorded during prior clinical care, findings from past OCT RNFL examinations (Carl Zeiss Cirrus, Dublin, CA), and findings from prior HFA3 or HFA2 SITA visual field tests. In a subsequent report, we will compare LC strain estimates for each eye with findings from prior RNFL and field testing.

### 2.5 Image selection and segmentation

Image volumes were selected, prioritizing optimal centering with a balance of clear focus, high contrast, and low noise, and imported into FIJI^31^ for post-processing. Contrast was enhanced to bring out the extracellular matrix (ECM) features of the LC using a contrast limited adaptive histogram equalization algorithm and Gamma filter in FIJI, as previously published.^21^ The post-processed image volumes were imported into MATLAB (R2024a, Mathworks, Natick, MA, US), and an automated segmentation algorithm was applied to identify the entire length of Bruch’s membrane and its end points at the ONH, the anterior border of the LC, and the choroidal—scleral interface.^32^ Each of the 24 scans was then inspected by a trained research coordinator to adjust the placement of these identified features, and their findings were validated using an inter-user variability test to ensure consistency. Higher order polynomial functions (MATLAB function *polyfit*) were fit to the (R,Z,Θ) pixel positions of the marked points of the tissue boundaries to segment the retina, choroid, sclera, and anterior LC border, as detailed in published work. In most images, the posterior limit of the LC was not identifiable, so the extent of the anterior LC region was defined by automatically offsetting the curve fitted to the anterior LC border posteriorly by 250 μm. The prelamina was defined as the tissue from the vitreoretinal interface to the LC border. The LC depth was measured from the line joining the two ends of Bruch’s membrane to the LC anterior border. For the present analysis, we present the LC strain data. Later analysis will include strain estimates for the retina, choroid, sclera, and prelamina.

### 2.6 LC strain estimation

LC strain was estimated by comparing the OCT volumes acquired pre- and post-medication change or SLT. DVC was used to determine the three-dimensional tissue displacement between imaging sessions, and the displacement gradient and strain tensor were subsequently calculated. The DVC and strain calculations were performed in a cylindrical coordinate system centered on the ONH to characterize radial, axial, and circumferential tissue deformation.

#### 2.6.1 Digital volume correlation

The Fast Iterative DVC (FI-DVC) algorithm, developed by Bar-Kochba et al.,^33^ was applied to compare the pre- and post-intervention images and calculate the three-dimensional deformation map in the imaged ONH volume. Briefly, the FI-DVC method determines the tissue deformation by iteratively tracking the speckle patterns present in the OCT volumes. Initially, the reference (pre-intervention) and deformed (post-intervention) images are divided into large overlapping subvolumes, and a Fast Fourier Transform implementation of the cross-correlation technique compares the patterns within each subvolume to estimate the first tissue displacement increment. The resulting deformation field is used to warp the images closer together, improving their alignment. In each subsequent iteration, the subvolume size is halved to improve the spatial resolution of the displacement correlation and accuracy of the displacement gradient and strain calculations, and the cross-correlation technique is then applied to the warped images to calculate the next displacement increment and the process is repeated until the displacement increment falls below a specified tolerance.

We modified the original FI-DVC algorithm to analyze radial SD-OCT scans in a cylindrical coordinate system centered on the ONH, calculating radial (*U_R_*), axial (*U_Z_*), and circumferential (*U_Θ_*) displacement fields as previously published.^21^ Complete displacement correlation across the ONH is not possible because of the blocking effect of overlying retinal blood vessels and variations in tissue reflectance. After applying displacement filters to exclude regions of poor correlation (or high correlation error), the percentage of the LC region with accurate DVC correlation was calculated for each eye by averaging across 24 radial scans. As in previous reports, eyes with DVC correlation in <20% of the LC volume were excluded from analysis.

#### 2.6.2 Strain calculation

After smoothing the displacement field as previously described, the displacement gradient and components of the Green-Lagrange strain tensor were calculated in the cylindrical coordinate system. The 6 strain components are radial strain (*E_rr_*), axial strain (*E_zz_*), circumferential strain (*E_θθ_*), and shear strains (*E_rz_*, *E_rθ_*, *E_zθ_*). For the normal strains (*E_rr_*, *E_θθ_, E_zz_*), negative strain indicates compression, and positive strain indicates expansion. The shear strain *E_zθ_* describes a twist about the z-axis. The maximum principal strain *E_max_* and maximum shear strain *Γ_max_* in the axial-radial (Z-R) plane were calculated as previously indicated.^21^ Effective strain,^25^ *E_eff_*, was calculated as:

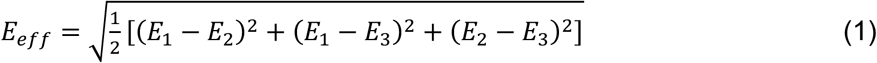

where *E_1_*, *E_2_*, and *E_3_* are the principal components of the Green-Lagrange strain tensor.

For each eye, DVC baseline (reproducibility) errors were estimated by applying the DVC algorithm to two image volumes acquired within short succession during the pre-intervention imaging session and calculating the differences in measured displacements and strains. DVC correlation errors were estimated by artificially warping an image: a rigid body translation was applied in the Z direction (*U_z_* = 10 μm) and uniform 2% and −2% strains were applied in the R and Z directions, respectively, before applying the DVC algorithm to assess the difference between the prescribed displacement and strain fields and those calculated by DVC.

### 2.7 Statistical analysis

Statistical analyses were performed using MATLAB (R2024a). Continuous patient-and eye-level demographic and clinical variables were summarized as mean ± standard deviation (SD) and median [interquartile range (IQR)]. Two-sample *t*-tests were used to compare continuous patient-level variables between participants who completed both pre- and post-intervention imaging and those who underwent pre-intervention imaging only. Discrete patient-level variables were compared between these groups using chi-squared tests. Eye-level variables were compared using linear mixed-effects models, with subject included as a random effect to account for the correlation between eyes from the same patient.

Potential outliers were identified using z-scores of strain components, with an eye excluded if it had an absolute z-score >5 for multiple strain components. Average strains, baseline errors, and correlation errors were summarized as mean ± SD and median [IQR] for the full dataset and for the subset of eyes with DVC correlation in >20% of the LC volume. Linear mixed-effects (LME) models were used to compare estimated values with zero strain and with baseline and correlation errors. For comparisons with error measurements, one-sided tests were used for strain components with a directional hypothesis based on the expected deformation of a cylinder (radial, circumferential, and axial strains), whereas two-sided tests were used for all other comparisons.

Associations between strain and IOP change were then examined to test the hypothesis that LC strain varies with the magnitude and direction of IOP change. Strain was the dependent variable and IOP change was the independent variable. Linear mixed-effects models were used for individual strain components, for which the direction of strain was expected to reverse with the direction of IOP change, as well as for effective strain. Quadratic mixed-effects models were used for maximum principal and shear strains to test the hypothesis that these measures, which characterize the magnitude of the deformation, increase with the magnitude of IOP change in either direction. Model selection was evaluated using the Akaike Information Criterion (AIC), with preference for the lowest-order model when AIC values were comparable.

Subsequent analyses examined whether demographic and clinical variables were associated with strain after accounting for IOP change. Each demographic or clinical variable was first evaluated individually in a model that also included IOP change. Variables with *p* < 0.10 in these individual adjusted models were retained as candidates for combined multivariable models, which were used to determine whether associations persisted when the candidate variables were considered together. This screening threshold was selected to reduce the likelihood of excluding potentially relevant variables during model development and was not used to define statistical significance.^34^

In all analyses, statistical significance was defined as *p* < 0.05.

## 3. Results

### 3.1 Study subject data

A total of 140 persons (242 eyes) underwent pre-intervention imaging. Of these, 205 eyes completed both pre- and post-intervention imaging; data for these eyes are presented in this report. The remaining 37 eyes from 21 persons were consented OAG patients who either declined temporary discontinuation of eyedrops and therefore did not receive post-intervention imaging (*n* = 32 eyes of 18 persons), or who did not complete post-intervention imaging after IOP change (*n* = 5 eyes of 3 persons). There were no significant differences between participants who did and did not complete post-intervention imaging, except for sex distribution, with proportionately more females among those who did not complete post-intervention imaging (Table 1).

**Table 1:**
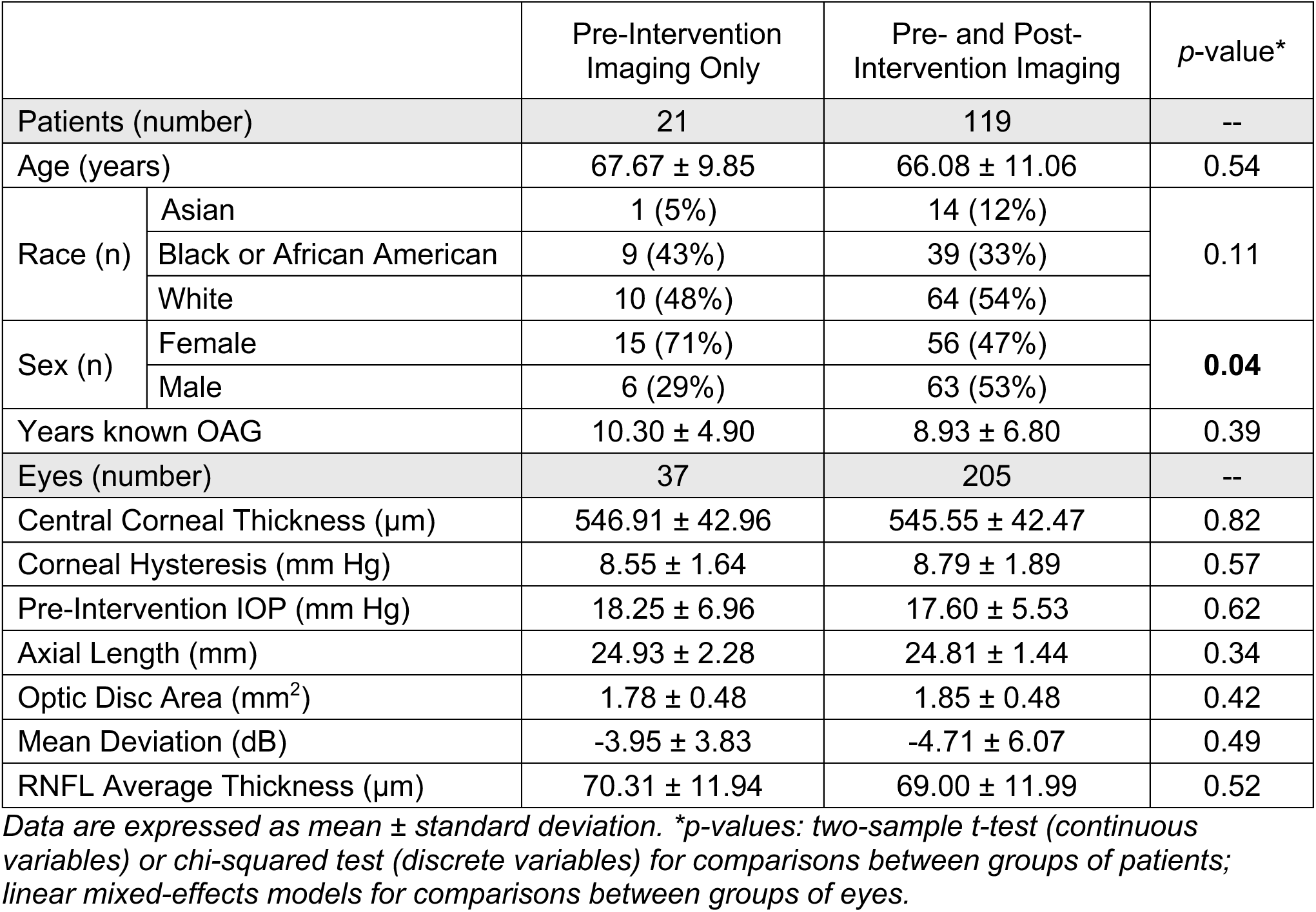
Demographics for subjects who received pre-intervention imaging only versus pre- and post-intervention imaging.

|  |  | Pre-Intervention Imaging Only | Pre- and Post-Intervention Imaging | *p*-value\* |
|---|---|---|---|---|
| Patients (number) |  | 21 | 119 | -- |
| Age (years) |  | 67.67 ± 9.85 | 66.08 ± 11.06 | 0.54 |
| Race (n) | Asian | 1 (5%) | 14 (12%) | 0.11 |
|  | Black or African American | 9 (43%) | 39 (33%) |  |
|  | White | 10 (48%) | 64 (54%) |  |
| Sex (n) | Female | 15 (71%) | 56 (47%) | **0.04** |
|  | Male | 6 (29%) | 63 (53%) |  |
| Years known OAG |  | 10.30 ± 4.90 | 8.93 ± 6.80 | 0.39 |
| Eyes (number) |  | 37 | 205 | -- |
| Central Corneal Thickness (µm) |  | 546.91 ± 42.96 | 545.55 ± 42.47 | 0.82 |
| Corneal Hysteresis (mm Hg) |  | 8.55 ± 1.64 | 8.79 ± 1.89 | 0.57 |
| Pre-Intervention IOP (mm Hg) |  | 18.25 ± 6.96 | 17.60 ± 5.53 | 0.62 |
| Axial Length (mm) |  | 24.93 ± 2.28 | 24.81 ± 1.44 | 0.34 |
| Optic Disc Area (mm<sup>2</sup>) |  | 1.78 ± 0.48 | 1.85 ± 0.48 | 0.42 |
| Mean Deviation (dB) |  | -3.95 ± 3.83 | -4.71 ± 6.07 | 0.49 |
| RNFL Average Thickness (µm) |  | 70.31 ± 11.94 | 69.00 ± 11.99 | 0.52 |

*Data are expressed as mean ± standard deviation. \*p-values: two-sample t-test (continuous variables) or chi-squared test (discrete variables) for comparisons between groups of patients; linear mixed-effects models for comparisons between groups of eyes.*

205 eyes of 119 subjects underwent post-intervention imaging after IOP change (Table 2), and DVC analysis was completed to estimate strains. The mean DVC correlation was 36.5 ± 18.1% of LC area (*n* = 205 eyes) and 42.0 ± 14.4% of LC area on a subset of 170 eyes with average DVC correlation > 20%, accounting for 83% of those with pre- and post-intervention image sets. Additional demographic and clinical data for the 205-eye dataset and 170-eye subset is summarized in Supplemental Table S1.

**Table 2:** Number of subjects included by category of examination and result.

|  | **# eyes** | **% eyes** | **# persons** | **% persons** |
|---|---|---|---|---|
| Completed DVC strain estimation | 205 | -- | 119 | -- |
| LC correlation > 20% | 170 | 83% | 105 | 88% |
| IOP change by temporary drop cessation (washout) | 153 | 75% | 83 | 70% |
| IOP change by starting new drop (wash-in) | 39 | 19% | 26 | 22% |
| IOP change by selective laser trabeculoplasty | 13 | 6% | 10 | 8% |

### 3.2 Medications added or selected for washout

The medications discontinued by washout patients, and those that were initiated by wash-in patients, are summarized in Supplemental Table S1. Across all patients, medications were predominately prostaglandin analogues, mostly latanoprost. Of 205 eyes, 109 were prescribed prostaglandin alone, and 10 eyes were prescribed prostaglandin in combination with other eye drops. Two of the 10 eyes were on 3 types of glaucoma drops. 56 eyes were receiving timolol, with 29 of these prescribed the dorzolamide-timolol combination. Four eyes were receiving dorzolamide, 3 used brimonidine, and 1 used netarsudil, all of these as single medications. The median time between change in treatment and post-intervention imaging across all eyes was 28 days.

### 3.3 Baseline and Correlation Error Data

Baseline displacement errors averaged over the correlated OCT volume in all 205 eyes were 0.004 ± 0.407 μm for *U_R_*, −0.035 ± 3.833 μm for *U_Θ_*, and −0.018 ± 0.258 μm for *U_Z_* in the washout group (*n* = 153, positive IOP change), and −0.043 ± 0.369 μm for *U_R_*, 0.083 ± 3.142 μm for *U_Θ_*, and −0.007 ± 0.244 μm for *U_Z_* in the wash-in/SLT group (*n* = 52, negative IOP change). This corresponded to less than 0.1 pixel error in the R-Z plane. Average baseline strain errors were significantly smaller than strains for *E_rr_* (0.029 ± 0.101%, *p* = 0.01) and *E_zz_* (0.060 ± 0.158%, *p* < 0.001) in the washout group, and for *E_rr_* (0.017 ± 0.093%, *p* = 0.05) and *E_zz_* (0.039 ± 0.126%, *p* = 0.001) in the wash-in/SLT group. The positive average *E_zz_* baseline strain errors across both groups indicated a slight tensile bias.

DVC correlation displacement errors averaged over the correlated OCT volume in all 205 eyes were −0.011 ± 0.073 μm for *U_R_*, −0.182 ± 0.351 μm for *U_Θ_*, and 0.620 ± 0.125 μm for *U_Z_* in the washout group, and −0.011 ± 0.105 μm for *U_R_*, −0.120 ± 0.356 μm for *U_Θ_*, and 0.623 ± 0.118 μm for *U_Z_* in the wash-in/SLT group. This corresponded to less than 1/5 pixel error in the R-Z plane. Average correlation strain errors were significantly smaller than strains for *E_rr_* −0.116 ± 0.066%, *p* < 0.001) and *E_zz_* (0.551 ± 0.076%, *p* < 0.001) in the washout group. Average *E_rr_* and *E_zz_* correlation errors in the wash-in/SLT group were similar to those in the washout group (*E_rr_* = −0.110 ± 0.067%, *p* = 0.65; *E_zz_* = 0.563 ± 0.089, *p* = 0.86).

Full results for average and absolute average baseline and correlation error are presented in Supplemental Tables S2-S3.

### 3.4 Strain data

We calculated average strains (individual strain components as well as maximum principal, maximum shear, and effective strains) for the overall LC (Figure 2), as well as for the four quadrants (superior, temporal, inferior, nasal). One eye with extreme strain values (z-score >5 for multiple strain components) was excluded from the strain analyses, resulting in a final analytical sample of 169 eyes of 104 persons. The mean and median pressures and average LC strains for these 169 eyes are shown in Table 3. LC strains for all 205 eyes are reported in Supplemental Table S4, and average LC strains by quadrant are reported in Supplemental Tables S5-S6. Pressures indicated here and in further analysis were measured in-office with iCare tonometer. Applanation measurements were used in eyes with missing iCare data (*n* = 45 of 205 eyes and *n* = 33 of the 169-eye subset).

**Figure 2:**
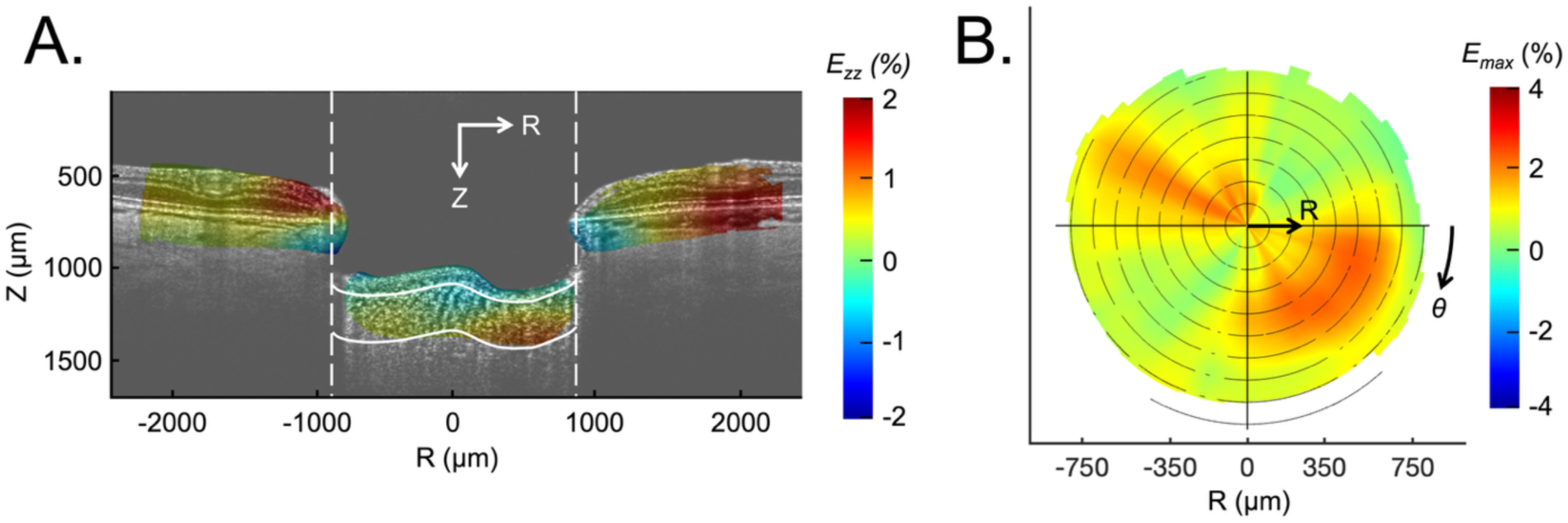
Example of strain field calculated using DVC. (A) Anterior-posterior strain (*E_zz_)* overlaid on the spectral domain OCT radial scan. (B) The *Z*-averaged maximum principal strain (*E_max_*) of the LC.

**Table 3:** Mean and median pressures and average LC strains.

|  | **Washout drop (*n* = 125)** |  |  | **New drop & SLT (*n* = 44)** |  |  |
|---|---|---|---|---|---|---|
|  | **Mean ± Std** | **Median [IQR]** | ***p*-value\*** | **Mean ± Std** | **Median [IQR]** | ***p*-value\*** |
| **Intraocular Pressure (mm Hg)** |  |  |  |  |  |  |
| **Pre-Intervention IOP** | 16.3 ± 4.8 | 16.0 [13.0, 18.3] | -- | 19.3 ± 5.2 | 19.0 [16.0, 22.5] | -- |
| **ΔIOP** | 2.9 ± 4.3 | 2.0 [0.0, 5.0] | -- | -4.3 ± 5.3 | -4.0 [-7.0, -3.0] | -- |
| **Average Strain (%)** |  |  |  |  |  |  |
| ***E<sub>rr</sub>*** | 0.081 ± 0.437 | 0.115 [-0.162, 0.298] | **0.05** | -0.051 ± 0.407 | -0.065 [-0.324, 0.174] | 0.40 |
| ***E<sub>θθ</sub>*** | -0.048 ± 0.843 | -0.009 [-0.503, 0.463] | 0.53 | -0.032 ± 1.130 | 0.009 [-0.786, 0.639] | 0.91 |
| ***E<sub>zz</sub>*** | -0.360 ± 0.798 | -0.293 [-0.668, 0.120] | **<0.001** | 0.398 ± 0.907 | 0.247 [-0.207, 0.669] | **0.02** |
| ***E<sub>rz</sub>*** | -0.194 ± 0.712 | -0.165 [-0.512, 0.203] | 0.51 | -0.011 ± 0.305 | -0.025 [-0.263, 0.153] | 0.81 |
| ***E<sub>rθ</sub>*** | 0.070 ± 0.656 | 0.118 [-0.258, 0.419] | **0.005** | 0.007 ± 0.630 | 0.002 [-0.286, 0.342] | 0.82 |
| ***E<sub>zθ</sub>*** | 0.913 ± 0.490 | 0.833 [0.537, 1.186] | 0.23 | 0.021 ± 0.705 | -0.097 [-0.413, 0.470] | 0.63 |
| ***E<sub>max</sub>*** | 1.052 ± 0.434 | 0.935 [0.754, 1.287] | **<0.001** | 1.242 ± 0.822 | 1.047 [0.718, 1.405] | **<0.001** |
| ***Γ<sub>max</sub>*** | 4.481 ± 1.994 | 4.009 [3.180, 5.096] | **<0.001** | 1.068 ± 0.440 | 0.955 [0.730, 1.356] | **<0.001** |
| ***E<sub>eff</sub>*** | 0.081 ± 0.437 | 0.115 [-0.162, 0.298] | **<0.001** | 4.792 ± 1.811 | 4.208 [3.529, 5.559] | **<0.001** |

*Std = standard deviation, IQR = interquartile range, ΔIOP = actual change in IOP. \*p-values: linear mixed-effects models vs. zero strain.*

Average LC strains for each eye were compared with IOP change, with positive values indicating an increase in IOP and negative values indicating a decrease in IOP. Axial strain (*E_zz_*) varied linearly with IOP change (*p* < 0.001), with increased IOP associated with more compressive (negative) strain and decreased IOP associated with more positive (tensile) strain (Figure 3). The relationship was approximately symmetric, with similar strain magnitudes observed for IOP increases and decreases of similar magnitude. Radial and circumferential strain also showed linear relationships with IOP change, although neither reached statistical significance, while the shear strain components were not significantly associated with IOP change.

**Figure 3:**
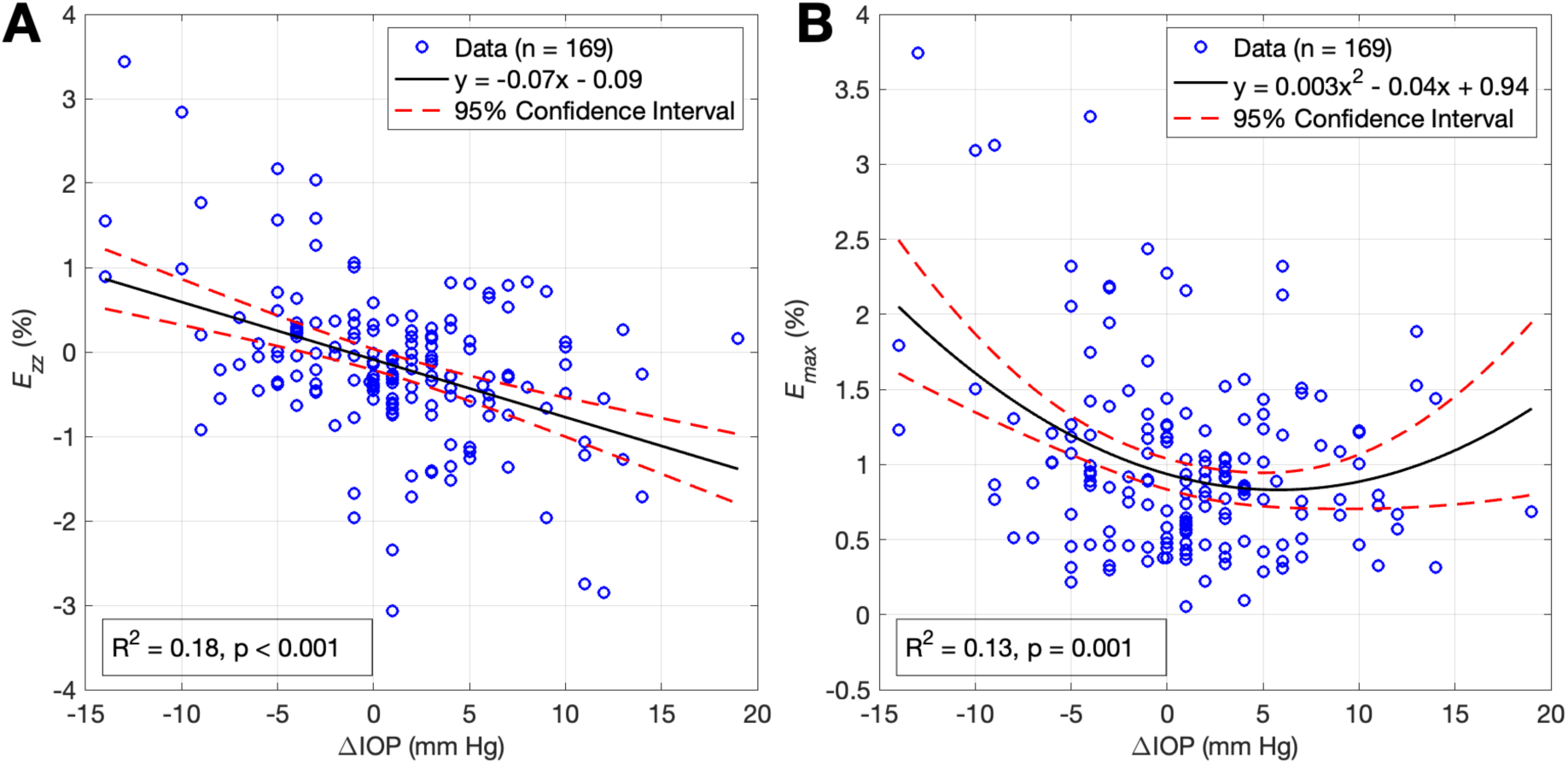
LC Strain versus IOP change. (A) Axial strain (*E_zz_*) showed a significant linear relationship with IOP change, with similar strain magnitudes observed for IOP increases and decreases of similar magnitude. (B) Maximum principal strain (*E_max_*) was positive for both IOP increases and decreases, and the quadratic term for its relationship with IOP change was significant. The *E_max_* relationship was asymmetric about zero IOP change.

Average maximum principal strain in the radial-axial plane (*E_max_*) was positive for both IOP increases and decreases and increased with the magnitude of IOP change. The quadratic term for the relationship between *E_max_* and IOP change was significant (*p* = 0.001), with some asymmetry about zero (Figure 3). Maximum shear strain in the radial-axial plane (*Γmax*) showed a similar relationship with IOP change, with a significant quadratic term (*p* = 0.02). Relationships between all average strains and IOP change are shown in Supplemental Figures S1-2.

### 3.5 Associations between strain and demographic/clinical variables

We constructed individual adjusted LME models to examine correlations between strain and demographic/clinical variables while adjusting for the degree and direction of IOP change. These relationships were modeled as:

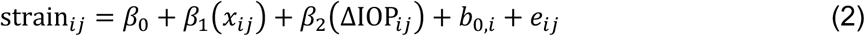

where strain*_i_*_j_, *x_i_*_j_, and ΔIOP*_i_*_j_ are the average strain, the demographic or clinical variable, and the change in IOP, respectively, for eye *j* of patient *i*. Additionally, *β*_0_ is the fixed intercept, *β*_1_ and *β*_2_ are the fixed effect coefficients, *b*_0,*i*_ is the random intercept for patient *i*, and *e_i_*_j_ is the residual error. For the six individual strain components, IOP change retained its directional sign: either positive (IOP increase) or negative (IOP decrease). For the maximum and effective strains (*E_max_*, *Γ_max_*, and *E_eff_*), which summarize the magnitude of the strain state and are consistently positive, models used the absolute value of IOP change to represent the magnitude of the pressure load.

The *p*-values for coefficient *β*_1_ (Equation 2) describing associations of demographic/clinical variable *x* with strain after adjusting for IOP change are summarized in Tables 4-5. Variables with significant relationships to one or more strains were RNFL thickness and its statistical abnormality grading (particularly inferiorly), age, race, axial length, and visual field MD. Lower RNFL thickness (Figure 4) and eyes with statistically abnormal RNFL thickness (Figure 5) were significantly associated with lower strains. Older age and longer axial length were also related to lower strains (Figures 6, 7). Visual field MD was related to circumferential strain *E_θθ_*, and Asian eyes had significantly greater shear strain *E_rz_* than those in the White reference group.

**Figure 4:**
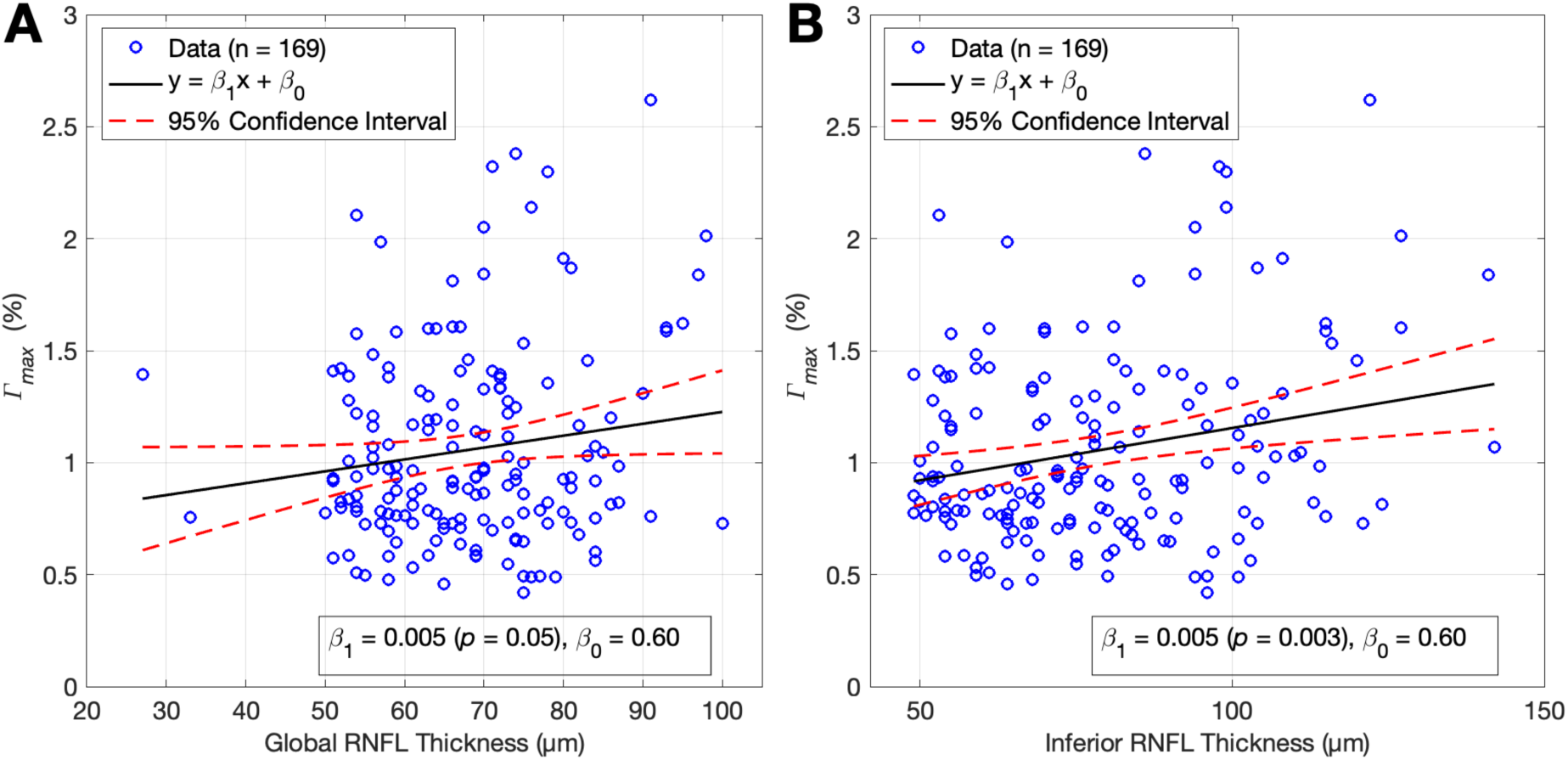
Maximum LC Shear Strain versus RNFL Thickness. (A) Average global RNFL thickness and (B) average inferior RNFL thickness each showed lower maximum shear strain with thinner RNFL. Fits represent the association between strain and RNFL thickness, adjusted for IOP change.

**Figure 5:**
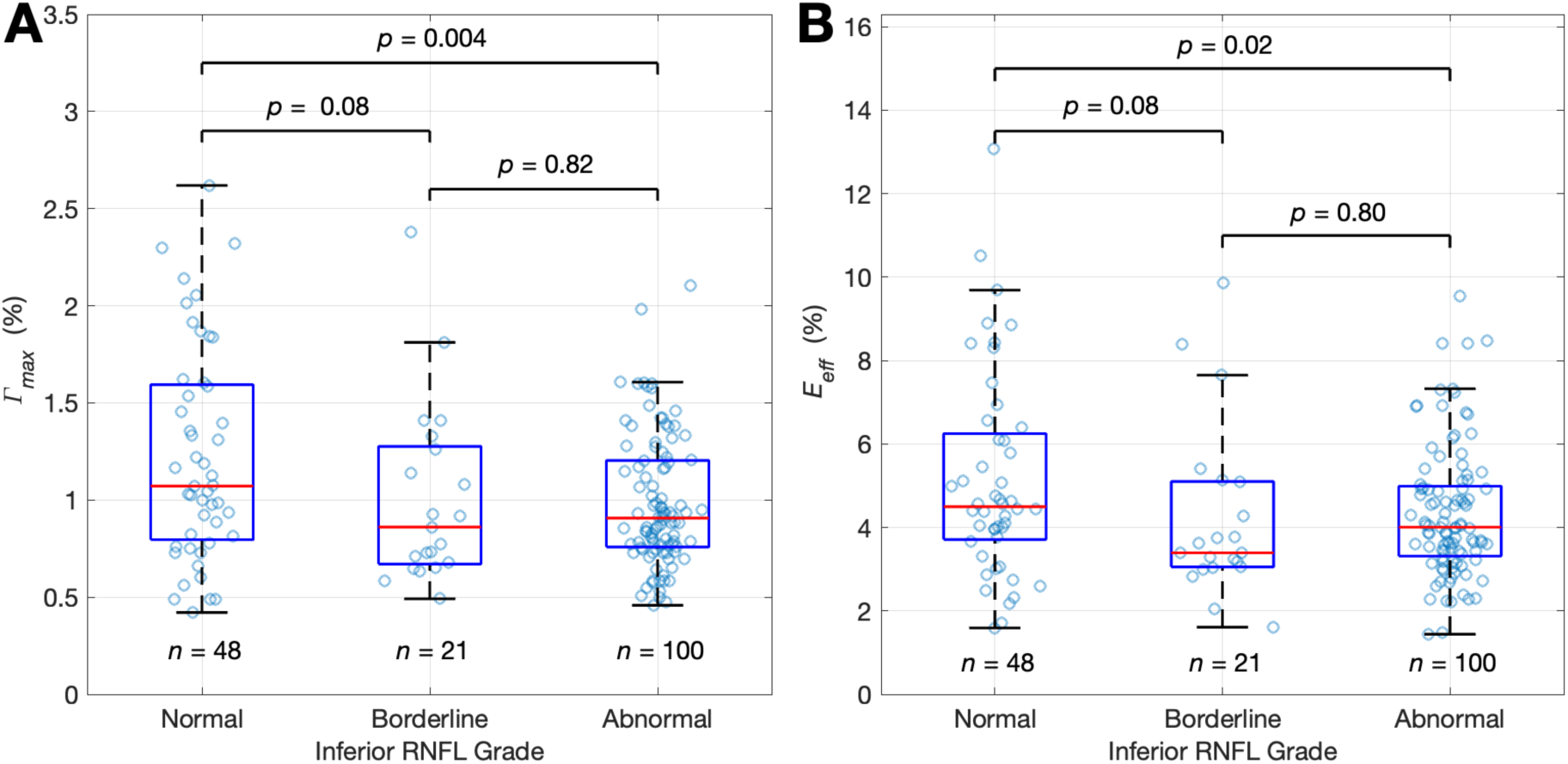
LC Strain versus Inferior RNFL Grade. Eyes with abnormal inferior RNFL grading had statistically lower (A) maximum shear strain (*Γ_max_*) and (B) effective strain (*E_eff_*) than eyes graded as normal. Eyes with borderline grading had the lowest strain values, though these differences did not reach statistical significance. Fits represent the association between strain and inferior RNFL grade, adjusted for IOP change.

**Figure 6:**
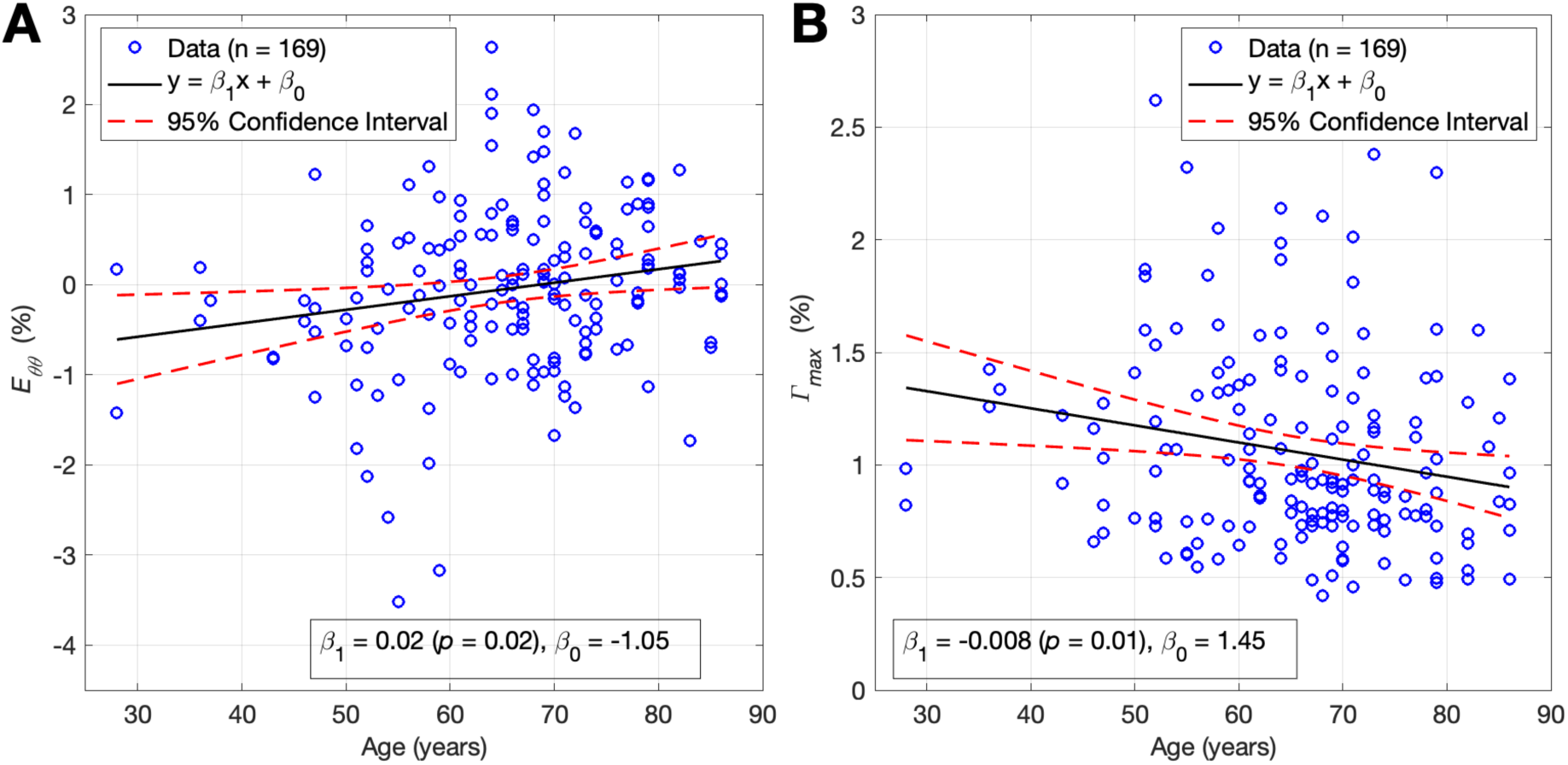
LC Strain versus Age. (A) Circumferential strain (*E_θθ_*) was greater with older age, while (B) maximum shear strain (*Γ_max_*) was lower in older eyes. Fits represent the association between strain and age, adjusted for IOP change.

**Figure 7:**
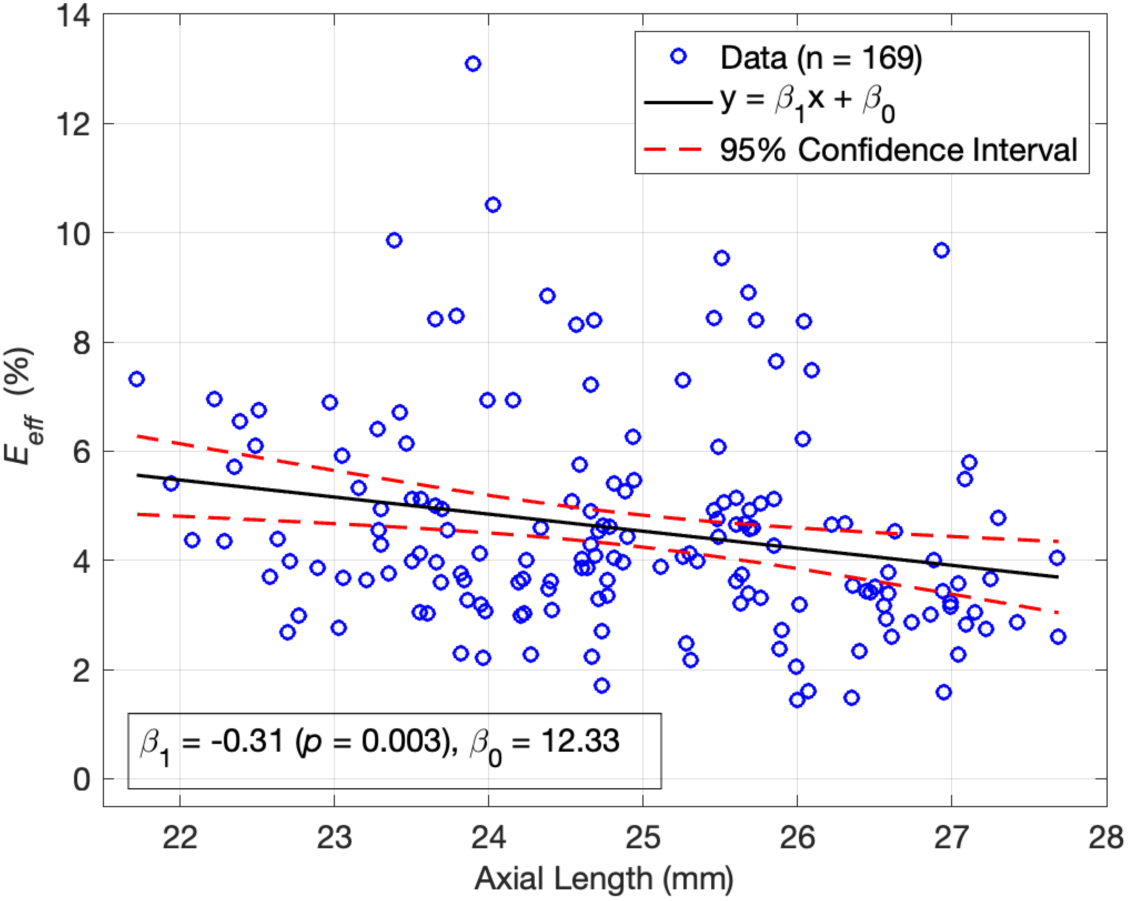
Effective LC Strain versus Axial Length. Longer eyes had lower effective strains. Fits represent the association between strain and axial length, adjusted for IOP change.

**Table 4:**
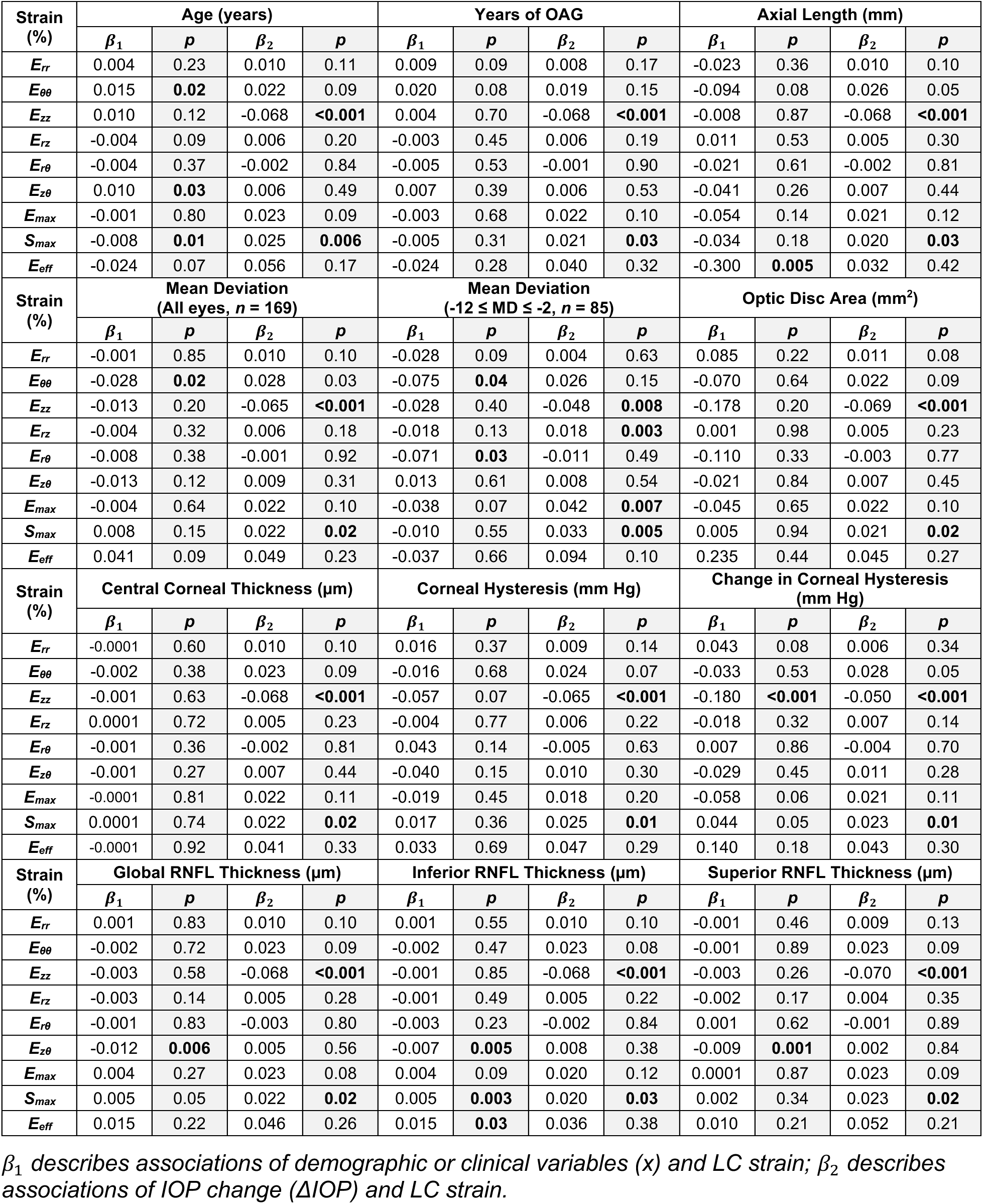
Coefficients and *p*-values for *β*_1_ and *β*_2_ in individual adjusted LME models with continuous demographic and clinical variables.

| **Strain (%)** | **Age (years)** |  |  |  | **Years of OAG** |  |  |  | **Axial Length (mm)** |  |  |  |
|---|---|---|---|---|---|---|---|---|---|---|---|---|
|  | ***β<sub>1</sub>*** | ***p*** | ***β<sub>2</sub>*** | ***p*** | ***β<sub>1</sub>*** | ***p*** | ***β<sub>2</sub>*** | ***p*** | ***β<sub>1</sub>*** | ***p*** | ***β<sub>2</sub>*** | ***p*** |
| ***E<sub>rr</sub>*** | 0.004 | 0.23 | 0.010 | 0.11 | 0.009 | 0.09 | 0.008 | 0.17 | -0.023 | 0.36 | 0.010 | 0.10 |
| ***E<sub>θθ</sub>*** | 0.015 | **0.02** | 0.022 | 0.09 | 0.020 | 0.08 | 0.019 | 0.15 | -0.094 | 0.08 | 0.026 | 0.05 |
| ***E<sub>zz</sub>*** | 0.010 | 0.12 | -0.068 | **<0.001** | 0.004 | 0.70 | -0.068 | **<0.001** | -0.008 | 0.87 | -0.068 | **<0.001** |
| ***E<sub>rz</sub>*** | -0.004 | 0.09 | 0.006 | 0.20 | -0.003 | 0.45 | 0.006 | 0.19 | 0.011 | 0.53 | 0.005 | 0.30 |
| ***E<sub>rθ</sub>*** | -0.004 | 0.37 | -0.002 | 0.84 | -0.005 | 0.53 | -0.001 | 0.90 | -0.021 | 0.61 | -0.002 | 0.81 |
| ***E<sub>zθ</sub>*** | 0.010 | **0.03** | 0.006 | 0.49 | 0.007 | 0.39 | 0.006 | 0.53 | -0.041 | 0.26 | 0.007 | 0.44 |
| ***E<sub>max</sub>*** | -0.001 | 0.80 | 0.023 | 0.09 | -0.003 | 0.68 | 0.022 | 0.10 | -0.054 | 0.14 | 0.021 | 0.12 |
| ***S<sub>max</sub>*** | -0.008 | **0.01** | 0.025 | **0.006** | -0.005 | 0.31 | 0.021 | **0.03** | -0.034 | 0.18 | 0.020 | **0.03** |
| ***E<sub>eff</sub>*** | -0.024 | 0.07 | 0.056 | 0.17 | -0.024 | 0.28 | 0.040 | 0.32 | -0.300 | **0.005** | 0.032 | 0.42 |
| **Strain (%)** | **Mean Deviation (All eyes, *n* = 169)** |  |  |  | **Mean Deviation (-12 ≤ MD ≤ -2, *n* = 85)** |  |  |  | **Optic Disc Area (mm<sup>2</sup>)** |  |  |  |
|  | ***β<sub>1</sub>*** | ***p*** | ***β<sub>2</sub>*** | ***p*** | ***β<sub>1</sub>*** | ***p*** | ***β<sub>2</sub>*** | ***p*** | ***β<sub>1</sub>*** | ***p*** | ***β<sub>2</sub>*** | ***p*** |
| ***E<sub>rr</sub>*** | -0.001 | 0.85 | 0.010 | 0.10 | -0.028 | 0.09 | 0.004 | 0.63 | 0.085 | 0.22 | 0.011 | 0.08 |
| ***E<sub>θθ</sub>*** | -0.028 | **0.02** | 0.028 | 0.03 | -0.075 | **0.04** | 0.026 | 0.15 | -0.070 | 0.64 | 0.022 | 0.09 |
| ***E<sub>zz</sub>*** | -0.013 | 0.20 | -0.065 | **<0.001** | -0.028 | 0.40 | -0.048 | **0.008** | -0.178 | 0.20 | -0.069 | **<0.001** |
| ***E<sub>rz</sub>*** | -0.004 | 0.32 | 0.006 | 0.18 | -0.018 | 0.13 | 0.018 | **0.003** | 0.001 | 0.98 | 0.005 | 0.23 |
| ***E<sub>rθ</sub>*** | -0.008 | 0.38 | -0.001 | 0.92 | -0.071 | **0.03** | -0.011 | 0.49 | -0.110 | 0.33 | -0.003 | 0.77 |
| ***E<sub>zθ</sub>*** | -0.013 | 0.12 | 0.009 | 0.31 | 0.013 | 0.61 | 0.008 | 0.54 | -0.021 | 0.84 | 0.007 | 0.45 |
| ***E<sub>max</sub>*** | -0.004 | 0.64 | 0.022 | 0.10 | -0.038 | 0.07 | 0.042 | **0.007** | -0.045 | 0.65 | 0.022 | 0.10 |
| ***S<sub>max</sub>*** | 0.008 | 0.15 | 0.022 | **0.02** | -0.010 | 0.55 | 0.033 | **0.005** | 0.005 | 0.94 | 0.021 | **0.02** |
| ***E<sub>eff</sub>*** | 0.041 | 0.09 | 0.049 | 0.23 | -0.037 | 0.66 | 0.094 | 0.10 | 0.235 | 0.44 | 0.045 | 0.27 |
| **Strain (%)** | **Central Corneal Thickness (µm)** |  |  |  | **Corneal Hysteresis (mm Hg)** |  |  |  | **Change in Corneal Hysteresis (mm Hg)** |  |  |  |
|  | ***β<sub>1</sub>*** | ***p*** | ***β<sub>2</sub>*** | ***p*** | ***β<sub>1</sub>*** | ***p*** | ***β<sub>2</sub>*** | ***p*** | ***β<sub>1</sub>*** | ***p*** | ***β<sub>2</sub>*** | ***p*** |
| ***E<sub>rr</sub>*** | -0.0001 | 0.60 | 0.010 | 0.10 | 0.016 | 0.37 | 0.009 | 0.14 | 0.043 | 0.08 | 0.006 | 0.34 |
| ***E<sub>θθ</sub>*** | -0.002 | 0.38 | 0.023 | 0.09 | -0.016 | 0.68 | 0.024 | 0.07 | -0.033 | 0.53 | 0.028 | 0.05 |
| ***E<sub>zz</sub>*** | -0.001 | 0.63 | -0.068 | **<0.001** | -0.057 | 0.07 | -0.065 | **<0.001** | -0.180 | **<0.001** | -0.050 | **<0.001** |
| ***E<sub>rz</sub>*** | 0.0001 | 0.72 | 0.005 | 0.23 | -0.004 | 0.77 | 0.006 | 0.22 | -0.018 | 0.32 | 0.007 | 0.14 |
| ***E<sub>rθ</sub>*** | -0.001 | 0.36 | -0.002 | 0.81 | 0.043 | 0.14 | -0.005 | 0.63 | 0.007 | 0.86 | -0.004 | 0.70 |
| ***E<sub>zθ</sub>*** | -0.001 | 0.27 | 0.007 | 0.44 | -0.040 | 0.15 | 0.010 | 0.30 | -0.029 | 0.45 | 0.011 | 0.28 |
| ***E<sub>max</sub>*** | -0.0001 | 0.81 | 0.022 | 0.11 | -0.019 | 0.45 | 0.018 | 0.20 | -0.058 | 0.06 | 0.021 | 0.11 |
| ***S<sub>max</sub>*** | 0.0001 | 0.74 | 0.022 | **0.02** | 0.017 | 0.36 | 0.025 | **0.01** | 0.044 | 0.05 | 0.023 | **0.01** |
| ***E<sub>eff</sub>*** | -0.0001 | 0.92 | 0.041 | 0.33 | 0.033 | 0.69 | 0.047 | 0.29 | 0.140 | 0.18 | 0.043 | 0.30 |
| **Strain (%)** | **Global RNFL Thickness (µm)** |  |  |  | **Inferior RNFL Thickness (µm)** |  |  |  | **Superior RNFL Thickness (µm)** |  |  |  |
|  | ***β<sub>1</sub>*** | ***p*** | ***β<sub>2</sub>*** | ***p*** | ***β<sub>1</sub>*** | ***p*** | ***β<sub>2</sub>*** | ***p*** | ***β<sub>1</sub>*** | ***p*** | ***β<sub>2</sub>*** | ***p*** |
| ***E<sub>rr</sub>*** | 0.001 | 0.83 | 0.010 | 0.10 | 0.001 | 0.55 | 0.010 | 0.10 | -0.001 | 0.46 | 0.009 | 0.13 |
| ***E<sub>θθ</sub>*** | -0.002 | 0.72 | 0.023 | 0.09 | -0.002 | 0.47 | 0.023 | 0.08 | -0.001 | 0.89 | 0.023 | 0.09 |
| ***E<sub>zz</sub>*** | -0.003 | 0.58 | -0.068 | **<0.001** | -0.001 | 0.85 | -0.068 | **<0.001** | -0.003 | 0.26 | -0.070 | **<0.001** |
| ***E<sub>rz</sub>*** | -0.003 | 0.14 | 0.005 | 0.28 | -0.001 | 0.49 | 0.005 | 0.22 | -0.002 | 0.17 | 0.004 | 0.35 |
| ***E<sub>rθ</sub>*** | -0.001 | 0.83 | -0.003 | 0.80 | -0.003 | 0.23 | -0.002 | 0.84 | 0.001 | 0.62 | -0.001 | 0.89 |
| ***E<sub>zθ</sub>*** | -0.012 | **0.006** | 0.005 | 0.56 | -0.007 | **0.005** | 0.008 | 0.38 | -0.009 | **0.001** | 0.002 | 0.84 |
| ***E<sub>max</sub>*** | 0.004 | 0.27 | 0.023 | 0.08 | 0.004 | 0.09 | 0.020 | 0.12 | 0.0001 | 0.87 | 0.023 | 0.09 |
| ***S<sub>max</sub>*** | 0.005 | 0.05 | 0.022 | **0.02** | 0.005 | **0.003** | 0.020 | **0.03** | 0.002 | 0.34 | 0.023 | **0.02** |
| ***E<sub>eff</sub>*** | 0.015 | 0.22 | 0.046 | 0.26 | 0.015 | **0.03** | 0.036 | 0.38 | 0.010 | 0.21 | 0.052 | 0.21 |

*β<sub>1</sub> describes associations of demographic or clinical variables (x) and LC strain; β<sub>2</sub> describes associations of IOP change (ΔIOP) and LC strain.*

**Table 5:**
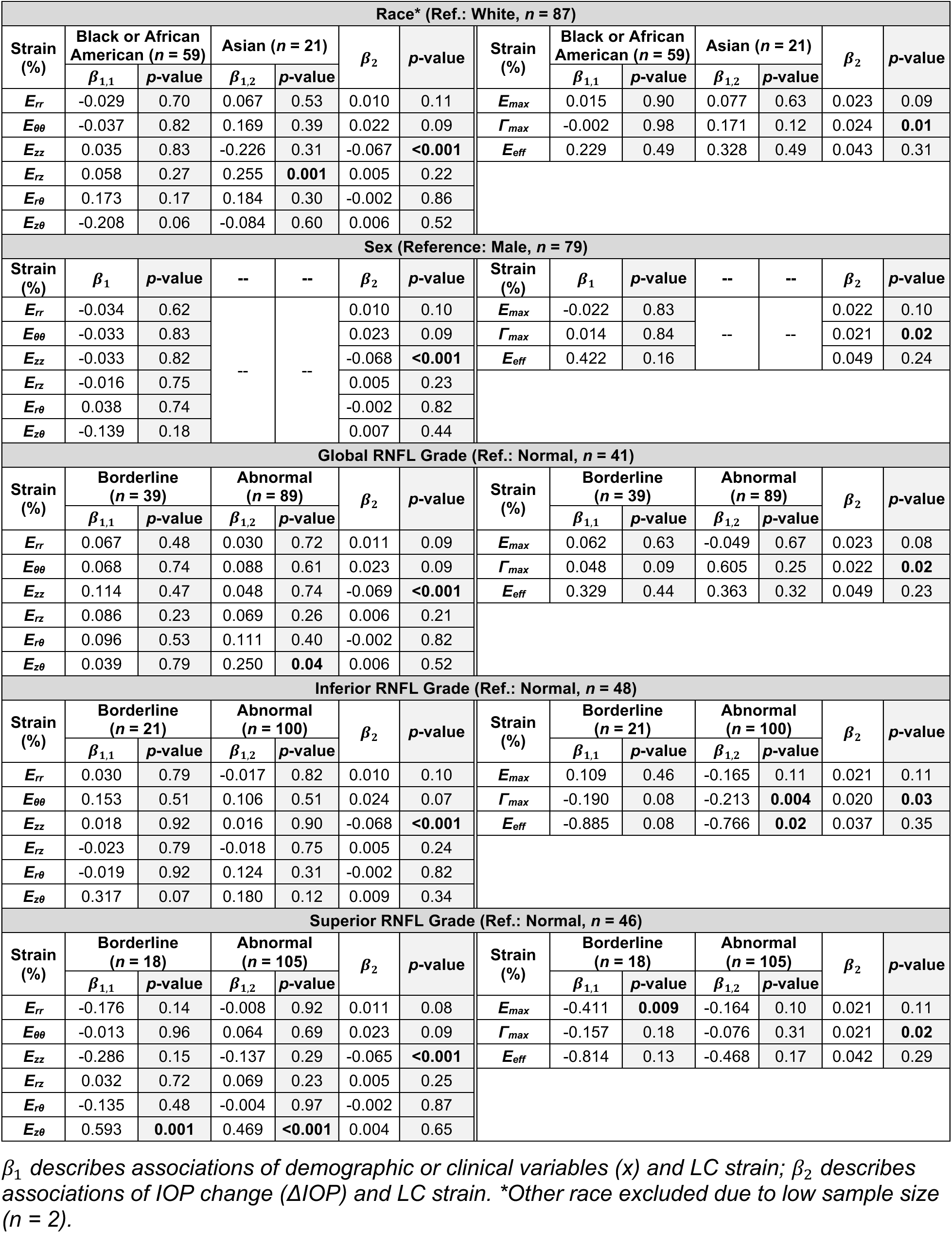
Coefficients and *p*-values for *β*_1_ and *β*_2_ in individual adjusted LME models with discrete demographic and clinical variables.

The values for sex, CCT, CH, years of known OAG, and disc area were not significantly associated with any strain after adjusting for IOP change. The change in CH between the pre- and post-intervention visits was significantly related to *E_zz_* strain, as will be discussed below and in a subsequent publication.

Across these models, coefficient *β*_2_ (Equation 2) describing associations of IOP change with strain was highly significant for axial strain *E_zz_* and significant for maximum shear strain *Γ_max_*, with the same directionality seen in Section 3.4 (Tables 4-5).

### 3.6 Associations between strain and multiple demographic/clinical variables

Having tested the relationships between strains and demographics variation and IOP, we next constructed combined multivariable LME models to simultaneously examine multiple demographic and clinical variables while continuing to adjust for the degree and direction of IOP change. These relationships were modeled as:

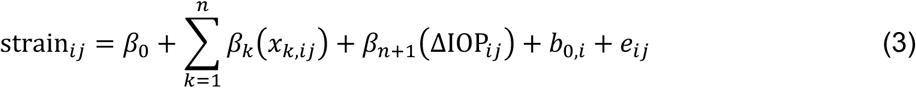

where *x_k_*_,*i*j_ represents demographic or clinical variable *k* for eye *j* of patient *i*. As before, strain*_i_*_j_and ΔIOP*_i_*_j_ are the average strain and the change in IOP, *β*_0_ is the fixed intercept, *β_k_* and *β_n_*_+1_ are the fixed effect coefficients, *b*_0,*i*_ is the random intercept for patient *i*, and *e_i_*_j_ is the residual error. The models again retained the directional sign of IOP change for the six individual strain components and used the absolute value of IOP change for the maximum and effective strains.

Demographic and clinical variables with *p* < 0.10 for at least one strain component in the individual adjusted models in Section 3.5 were considered for the combined models. In addition to IOP change, these included age, race, axial length, and either visual field MD or RNFL thickness (Table 6). In every model, IOP change remained significantly associated with axial strain *E_zz_* (*p* < 0.001), with the same directionality seen in Section 3.4. Average global, inferior, and superior quadrant RNFL thicknesses were associated with shear strain component *E_zθ_* (*p* ≤ 0.006). Inferior RNFL thickness was additionally associated with maximum shear strain *Γ_max_* (*p* = 0.02), with lower RNFL thickness associated with lower strain. Age and axial length were significantly associated with maximum shear strain *Γ_max_* and effective strain *E_eff_* in all models, with older age and larger axial length associated with lower strain. Age was additionally related to circumferential strain *E_θθ_*, and eyes from participants who identified as Asian had significantly greater shear strain *E_rz_* than those from participants who identified as White (reference group). Visual field MD had no significant relationships to strain in these models.

**Table 6:**
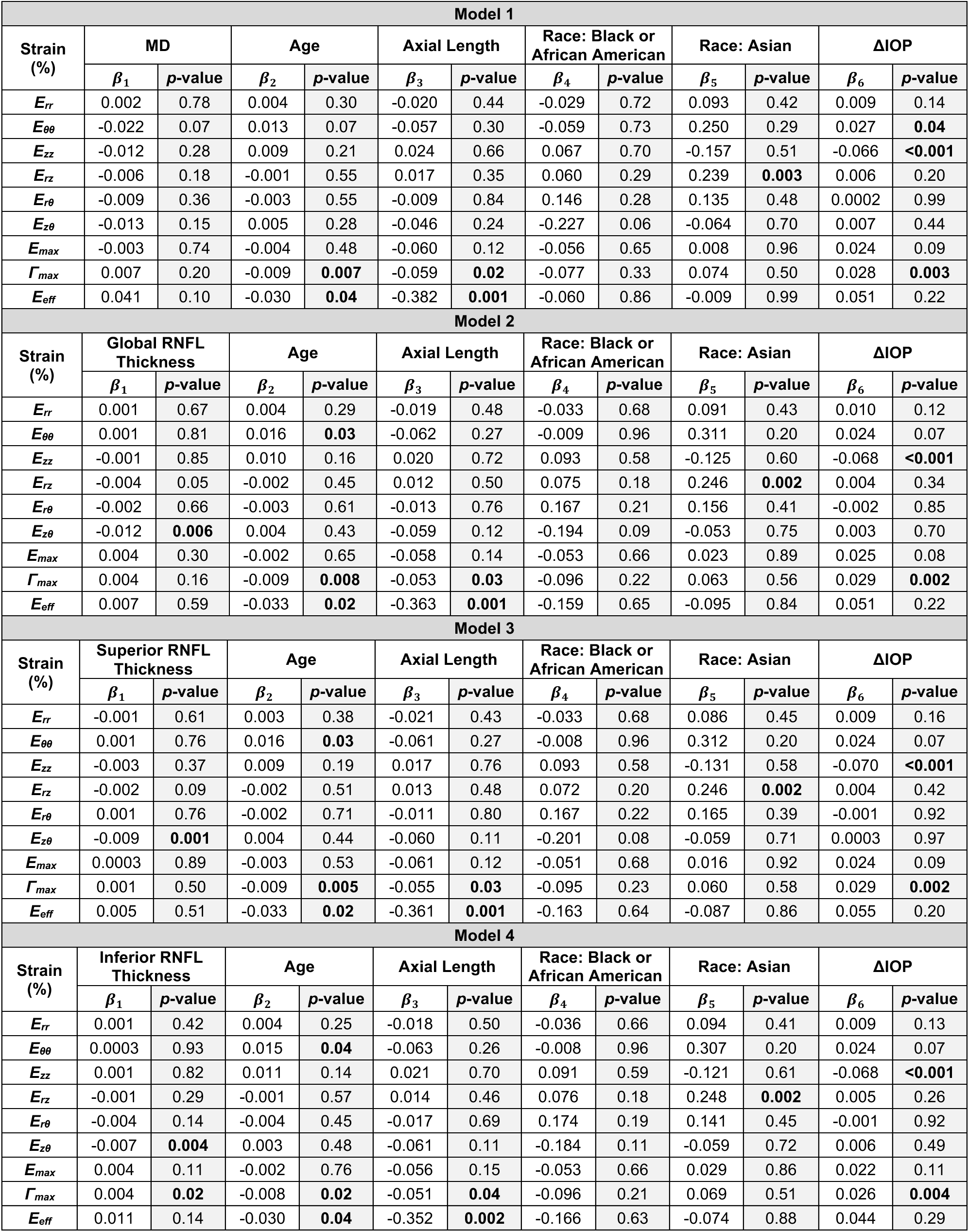
Coefficients and *p*-values for multivariable LME models including multiple demographic and clinical variables.

| **Model 1** |  |  |  |  |  |  |  |  |  |  |  |  |
|---|---|---|---|---|---|---|---|---|---|---|---|---|
| **Strain (%)** | **MD** |  | **Age** |  | **Axial Length** |  | **Race: Black or African American** |  | **Race: Asian** |  | **ΔIOP** |  |
|  | ***β<sub>1</sub>*** | ***p*-value** | ***β<sub>2</sub>*** | ***p*-value** | ***β<sub>3</sub>*** | ***p*-value** | ***β<sub>4</sub>*** | ***p*-value** | ***β<sub>5</sub>*** | ***p*-value** | ***β<sub>6</sub>*** | ***p*-value** |
| ***E<sub>rr</sub>*** | 0.002 | 0.78 | 0.004 | 0.30 | -0.020 | 0.44 | -0.029 | 0.72 | 0.093 | 0.42 | 0.009 | 0.14 |
| ***E<sub>θθ</sub>*** | -0.022 | 0.07 | 0.013 | 0.07 | -0.057 | 0.30 | -0.059 | 0.73 | 0.250 | 0.29 | 0.027 | **0.04** |
| ***E<sub>zz</sub>*** | -0.012 | 0.28 | 0.009 | 0.21 | 0.024 | 0.66 | 0.067 | 0.70 | -0.157 | 0.51 | -0.066 | **<0.001** |
| ***E<sub>rz</sub>*** | -0.006 | 0.18 | -0.001 | 0.55 | 0.017 | 0.35 | 0.060 | 0.29 | 0.239 | **0.003** | 0.006 | 0.20 |
| ***E<sub>rθ</sub>*** | -0.009 | 0.36 | -0.003 | 0.55 | -0.009 | 0.84 | 0.146 | 0.28 | 0.135 | 0.48 | 0.0002 | 0.99 |
| ***E<sub>zθ</sub>*** | -0.013 | 0.15 | 0.005 | 0.28 | -0.046 | 0.24 | -0.227 | 0.06 | -0.064 | 0.70 | 0.007 | 0.44 |
| ***E<sub>max</sub>*** | -0.003 | 0.74 | -0.004 | 0.48 | -0.060 | 0.12 | -0.056 | 0.65 | 0.008 | 0.96 | 0.024 | 0.09 |
| ***Γ<sub>max</sub>*** | 0.007 | 0.20 | -0.009 | **0.007** | -0.059 | **0.02** | -0.077 | 0.33 | 0.074 | 0.50 | 0.028 | **0.003** |
| ***E<sub>eff</sub>*** | 0.041 | 0.10 | -0.030 | **0.04** | -0.382 | **0.001** | -0.060 | 0.86 | -0.009 | 0.99 | 0.051 | 0.22 |
| **Model 2** |  |  |  |  |  |  |  |  |  |  |  |  |
| **Strain (%)** | **Global RNFL Thickness** |  | **Age** |  | **Axial Length** |  | **Race: Black or African American** |  | **Race: Asian** |  | **ΔIOP** |  |
|  | ***β<sub>1</sub>*** | ***p*-value** | ***β<sub>2</sub>*** | ***p*-value** | ***β<sub>3</sub>*** | ***p*-value** | ***β<sub>4</sub>*** | ***p*-value** | ***β<sub>5</sub>*** | ***p*-value** | ***β<sub>6</sub>*** | ***p*-value** |
| ***E<sub>rr</sub>*** | 0.001 | 0.67 | 0.004 | 0.29 | -0.019 | 0.48 | -0.033 | 0.68 | 0.091 | 0.43 | 0.010 | 0.12 |
| ***E<sub>θθ</sub>*** | 0.001 | 0.81 | 0.016 | **0.03** | -0.062 | 0.27 | -0.009 | 0.96 | 0.311 | 0.20 | 0.024 | 0.07 |
| ***E<sub>zz</sub>*** | -0.001 | 0.85 | 0.010 | 0.16 | 0.020 | 0.72 | 0.093 | 0.58 | -0.125 | 0.60 | -0.068 | **<0.001** |
| ***E<sub>rz</sub>*** | -0.004 | 0.05 | -0.002 | 0.45 | 0.012 | 0.50 | 0.075 | 0.18 | 0.246 | **0.002** | 0.004 | 0.34 |
| ***E<sub>rθ</sub>*** | -0.002 | 0.66 | -0.003 | 0.61 | -0.013 | 0.76 | 0.167 | 0.21 | 0.156 | 0.41 | -0.002 | 0.85 |
| ***E<sub>zθ</sub>*** | -0.012 | **0.006** | 0.004 | 0.43 | -0.059 | 0.12 | -0.194 | 0.09 | -0.053 | 0.75 | 0.003 | 0.70 |
| ***E<sub>max</sub>*** | 0.004 | 0.30 | -0.002 | 0.65 | -0.058 | 0.14 | -0.053 | 0.66 | 0.023 | 0.89 | 0.025 | 0.08 |
| ***Γ<sub>max</sub>*** | 0.004 | 0.16 | -0.009 | **0.008** | -0.053 | **0.03** | -0.096 | 0.22 | 0.063 | 0.56 | 0.029 | **0.002** |
| ***E<sub>eff</sub>*** | 0.007 | 0.59 | -0.033 | **0.02** | -0.363 | **0.001** | -0.159 | 0.65 | -0.095 | 0.84 | 0.051 | 0.22 |
| **Model 3** |  |  |  |  |  |  |  |  |  |  |  |  |
| **Strain (%)** | **Superior RNFL Thickness** |  | **Age** |  | **Axial Length** |  | **Race: Black or African American** |  | **Race: Asian** |  | **ΔIOP** |  |
|  | ***β<sub>1</sub>*** | ***p*-value** | ***β<sub>2</sub>*** | ***p*-value** | ***β<sub>3</sub>*** | ***p*-value** | ***β<sub>4</sub>*** | ***p*-value** | ***β<sub>5</sub>*** | ***p*-value** | ***β<sub>6</sub>*** | ***p*-value** |
| ***E<sub>rr</sub>*** | -0.001 | 0.61 | 0.003 | 0.38 | -0.021 | 0.43 | -0.033 | 0.68 | 0.086 | 0.45 | 0.009 | 0.16 |
| ***E<sub>θθ</sub>*** | 0.001 | 0.76 | 0.016 | **0.03** | -0.061 | 0.27 | -0.008 | 0.96 | 0.312 | 0.20 | 0.024 | 0.07 |
| ***E<sub>zz</sub>*** | -0.003 | 0.37 | 0.009 | 0.19 | 0.017 | 0.76 | 0.093 | 0.58 | -0.131 | 0.58 | -0.070 | **<0.001** |
| ***E<sub>rz</sub>*** | -0.002 | 0.09 | -0.002 | 0.51 | 0.013 | 0.48 | 0.072 | 0.20 | 0.246 | **0.002** | 0.004 | 0.42 |
| ***E<sub>rθ</sub>*** | 0.001 | 0.76 | -0.002 | 0.71 | -0.011 | 0.80 | 0.167 | 0.22 | 0.165 | 0.39 | -0.001 | 0.92 |
| ***E<sub>zθ</sub>*** | -0.009 | **0.001** | 0.004 | 0.44 | -0.060 | 0.11 | -0.201 | 0.08 | -0.059 | 0.71 | 0.0003 | 0.97 |
| ***E<sub>max</sub>*** | 0.0003 | 0.89 | -0.003 | 0.53 | -0.061 | 0.12 | -0.051 | 0.68 | 0.016 | 0.92 | 0.024 | 0.09 |
| ***Γ<sub>max</sub>*** | 0.001 | 0.50 | -0.009 | **0.005** | -0.055 | **0.03** | -0.095 | 0.23 | 0.060 | 0.58 | 0.029 | **0.002** |
| ***E<sub>eff</sub>*** | 0.005 | 0.51 | -0.033 | **0.02** | -0.361 | **0.001** | -0.163 | 0.64 | -0.087 | 0.86 | 0.055 | 0.20 |
| **Model 4** |  |  |  |  |  |  |  |  |  |  |  |  |
| **Strain (%)** | **Inferior RNFL Thickness** |  | **Age** |  | **Axial Length** |  | **Race: Black or African American** |  | **Race: Asian** |  | **ΔIOP** |  |
|  | ***β<sub>1</sub>*** | ***p*-value** | ***β<sub>2</sub>*** | ***p*-value** | ***β<sub>3</sub>*** | ***p*-value** | ***β<sub>4</sub>*** | ***p*-value** | ***β<sub>5</sub>*** | ***p*-value** | ***β<sub>6</sub>*** | ***p*-value** |
| ***E<sub>rr</sub>*** | 0.001 | 0.42 | 0.004 | 0.25 | -0.018 | 0.50 | -0.036 | 0.66 | 0.094 | 0.41 | 0.009 | 0.13 |
| ***E<sub>θθ</sub>*** | 0.0003 | 0.93 | 0.015 | **0.04** | -0.063 | 0.26 | -0.008 | 0.96 | 0.307 | 0.20 | 0.024 | 0.07 |
| ***E<sub>zz</sub>*** | 0.001 | 0.82 | 0.011 | 0.14 | 0.021 | 0.70 | 0.091 | 0.59 | -0.121 | 0.61 | -0.068 | **<0.001** |
| ***E<sub>rz</sub>*** | -0.001 | 0.29 | -0.001 | 0.57 | 0.014 | 0.46 | 0.076 | 0.18 | 0.248 | **0.002** | 0.005 | 0.26 |
| ***E<sub>rθ</sub>*** | -0.004 | 0.14 | -0.004 | 0.45 | -0.017 | 0.69 | 0.174 | 0.19 | 0.141 | 0.45 | -0.001 | 0.92 |
| ***E<sub>zθ</sub>*** | -0.007 | **0.004** | 0.003 | 0.48 | -0.061 | 0.11 | -0.184 | 0.11 | -0.059 | 0.72 | 0.006 | 0.49 |
| ***E<sub>max</sub>*** | 0.004 | 0.11 | -0.002 | 0.76 | -0.056 | 0.15 | -0.053 | 0.66 | 0.029 | 0.86 | 0.022 | 0.11 |
| ***Γ<sub>max</sub>*** | 0.004 | **0.02** | -0.008 | **0.02** | -0.051 | **0.04** | -0.096 | 0.21 | 0.069 | 0.51 | 0.026 | **0.004** |
| ***E<sub>eff</sub>*** | 0.011 | 0.14 | -0.030 | **0.04** | -0.352 | **0.002** | -0.166 | 0.63 | -0.074 | 0.88 | 0.044 | 0.29 |

## 4. Discussion

The STRAIN study is a prospective clinical trial investigating LC strain as a potential prognostic biomarker in OAG, and here, we present an initial cross-sectional analysis of baseline data. We used OCT imaging and DVC to estimate LC strain in a large cohort of OAG patients following IOP alteration induced by changes in glaucoma medication or SLT. LC strain varied with the magnitude and direction of IOP change, and several strain measures were associated with demographic and ocular characteristics, including age, race, axial length, and RNFL thickness. In the following sections, we discuss the LC strain response to IOP change, its associations with glaucoma damage and other clinical characteristics, and the potential implications of these findings for the use of LC strain as a prognostic factor in OAG.

### 4.1 LC strain varies with magnitude and direction of IOP change

The level of IOP is an established risk and prognostic factor for OAG.^2,3^ Previous studies have assessed LC strain following both IOP elevation and IOP reduction, but the methods used to alter IOP generally differed between the two directions. IOP elevation has been induced acutely using ophthalmodynamometry^26,28,29^ or tight swimming goggles,^23^ whereas IOP reduction has been studied following trabeculectomy,^25^ suture lysis after trabeculectomy,^22^ or initiation of glaucoma eye drops.^24^ LC strain has also been estimated following alteration of intracranial pressure (ICP).^35^ Thus, previous studies have differed in the direction, magnitude, and method of IOP alteration, as well as the duration over which the pressure change was maintained.

We estimated LC strain following both IOP increase and IOP decrease using the same clinical framework for IOP alteration, imaging, and strain analysis. This approach allowed us to examine the LC strain response to IOP change in both directions using a consistent protocol. We found that greater changes in IOP were associated with greater strain magnitudes. Among the individual strain components, axial strain was the largest in magnitude and demonstrated a significant linear relationship with IOP change, such that IOP increases and decreases of similar magnitude were associated with strain responses of similar magnitude but opposite sign. The direction of this association is consistent with prior studies demonstrating tensile axial LC strain in response to IOP reduction by suture lysis and compressive axial LC strain in response to IOP elevation by tight-fitting goggles; however, strain responses following these two methods of IOP alteration were not consistently symmetric.^22,23^ Prior work has attributed this asymmetry to the fact that compression-induced IOP elevation using approaches such as goggles or ophthalmodynamometry acts partly through orbital compression rather than through changes in aqueous humor dynamics, which may complicate comparisons between compression-based and other methods of IOP alteration. Our use of a consistent, non-mechanical method of IOP alteration in both directions may contribute to the more symmetric axial strain response observed here. Magnitudes of the remaining strain components were smaller than that of axial strain. Radial and circumferential strain also showed linear relationships with IOP change, though these relationships did not reach statistical significance, and the shear strain components were not associated with IOP change. These differing relationships among strain components suggest a complex LC strain response to IOP change. The multidirectional LC response may also depend on changes in pressure across the LC, as we previously found that individual strain components differed in their sensitivity to changes in IOP and ICP.^35^

In contrast, maximum principal strain, which indicates the maximum normal strain in the axial-radial plane, was positive for both IOP elevation and reduction, and the quadratic term for its relationship with IOP change was significant. The association showed some asymmetry around zero IOP change, which may reflect differences in the principal strain direction between IOP elevation and reduction. Together with the methodological variability among prior studies noted above, these results highlight the importance of considering the method and duration of IOP alteration, as well as the specific strain component or norm examined, when comparing LC strain responses across studies.

### 4.2 LC strain varies with age and OAG damage

In addition to higher IOP, older age is a consistent risk and prognostic factor for OAG. We found that LC circumferential strain and circumferential-axial shear strain, as well as maximum shear strain, were lower in older OAG eyes, consistent with previous observations of lower LC strain with age.^17^ Associations of age with circumferential and maximum shear strain remained significant in the combined multivariable models. Previous histological and other experimental studies suggest that age-related changes in LC composition and architecture may contribute to the lower strains observed in older eyes. The LC consists of approximately 50% collagen, 25% elastin,^36^ and ∼2% proteoglycans in dry weight,^37,38^ and histological studies have shown that LC fiber content increases with age. An increase in LC fiber content may contribute to thicker LC beams, which are associated with smaller strains.^16,39^ Other components of the LC extracellular matrix may also influence its mechanical response. Experimental removal of sulfated glycosaminoglycans led to decreased LC strain in *postmortem* eyes, with greater decreases in strain in eyes from younger individuals and in regions with larger baseline strain magnitudes.^40^ However, while the effect of age alone on LC composition is compatible with a decrease in LC strain,^41^ damage and remodeling in glaucoma may lead to structural changes that also impact the mechanical response of the LC. Histological analysis of OAG eyes has found decreased collagen density and altered elastin organization within LC beams compared to age-matched eyes, with elastin fibers becoming increasingly curved with greater glaucomatous damage.^42,43^ LC beam width was thinner in eyes with greater axon loss.^10^

In a small number of eyes (*n* = 17), Hannay et al.^24^ found that axial, maximum principal, and maximum shear strains were larger with more abnormal visual field MD. Czerpak et al.^22^ also found for a study of *n* = 26 eyes a more compliant *E_max_, Γ_max_*, and *E_rθ_* strain response to IOP change for eyes with worse, more negative MD. In the larger sample and wider range of MD values in the present study, however, eyes with worse MD had slightly lower maximum principal and maximum shear strains, though this association was not significant. The relationship between glaucomatous neural damage and visual field MD is nonlinear, with relatively small changes in MD accompanying retinal ganglion cell loss in early disease.^44^ The present cohort was weighted toward mild visual field loss, with an average MD of −4.71 ± 6.7 dB, compared to the cohort in Hannay et al. (−6.6 ± 5.8 dB) with nearly equal distribution of severe/moderate (MD < −6 dB) and mild damage (MD < −6 dB) eyes, and in Czerpak (−11.5 ± 5.8 dB) where two thirds of eyes had moderate/severe damage. The limited sensitivity of MD to neural damage in early disease may have reduced the ability to detect an association with LC strain in this study.

In contrast, LC strain was associated with structural measures of glaucomatous damage. One recent study found that strain-sensitive regions of the ONH corresponded spatially with regions associated with visual field loss.^45^ In our study, decreased RNFL thickness was associated with decreased strain, particularly shear strain in the inferior quadrant. Associations between RNFL thickness and shear strain remained significant in the combined multivariable models, including associations of inferior RNFL thickness with maximum shear strain. The stronger association between inferior LC strain and inferior RNFL thickness loss is consistent with the known greater susceptibility of the inferior quadrant of the ONH to glaucoma damage. When eyes were examined by RNFL grading, those with abnormal RNFL thickness had significantly lower maximum shear and effective strain in the inferior quadrant than those with normal RNFL thickness. In the present study, eyes with borderline RNFL thickness exhibited the lowest LC strain of the normal, borderline, and abnormal groups, although this difference was not statistically significant, potentially reflecting the smaller number of eyes in this group. A similar nonmonotonic pattern was found in *ex vivo* inflation testing of human donor eyes with glaucoma.^19^ One possible explanation for this pattern is that ONH remodeling, a dynamic process that evolves with disease progression, alters the mechanical response of the LC.^12^ Hannay et al.^24^ also found a lower strain response with thinner RNFL, while Czerpak et al.^22^ reported the opposite trend. The eyes studied by Czerpak et al. had a higher proportion of moderate/severe disease than those in Hannay et al. and this study. These differing trends support a nonmonotonic relationship between RNFL thickness and LC strain, where a stiffer strain response is associated with greater axon loss in early glaucoma, while a more compliant strain response is associated with worse axon loss in late glaucoma. Changes in the ONH mechanical response with disease progression have also been observed in experimental glaucoma, with increased compliance detected early after glaucoma induction that subsequently decreased toward that of control eyes.^46^ This possibility is further supported by our previous finding that LC strain continued to change for months following IOP change, consistent with remodeling of the LC.^47^ The pattern across RNFL grades observed here may therefore reflect the interplay between structural changes and remodeling in glaucoma. Differences in LC strain may reflect both biomechanical properties associated with susceptibility to axonal injury and alterations in LC mechanical behavior resulting from glaucomatous remodeling.

Together, these findings suggest that both age and glaucomatous damage may alter LC structure and, consequently, influence LC strain. Their effects may not necessarily be in the same direction or arise through the same mechanisms. Therefore, both age and the extent of glaucomatous damage should be considered when interpreting LC strain in relation to OAG progression. We expect that longitudinal data from the STRAIN study will help to clarify the association between LC strain and progressive OAG disease.

### 4.3 Additional parameters are associated with LC strain

We found that longer axial length, as occurs in axial myopia, was associated with lower LC effective strain. This association remained significant in the combined multivariable models when other demographic and clinical variables were considered simultaneously. This differs from our previous finding that longer axial length was associated with greater LC strain following acute IOP increase and decrease,^22,23^ potentially reflecting differences in the LC response to acute versus sustained IOP changes. Chuangsuwanich et al. similarly examined the relationship between myopia and LC effective strain, reporting greater LC effective strain in highly myopic eyes with OAG than highly myopic eyes without glaucoma following acute IOP elevation, and no difference in effective strain between non-myopic and highly myopic controls.^48^ Although the different study designs preclude a direct comparison, these findings raise questions about whether the association between axial length and LC strain varies across populations or glaucoma severity. In particular, the prior study consisted of a Chinese cohort, whereas our OAG cohort included participants from diverse racial backgrounds, and radial-axial shear strain was greater in eyes from participants who identified as Asian than those who identified as White in the combined multivariable models. These findings warrant further investigation, particularly given differences in the distribution of IOP among Asian and European populations with OAG.

Thinner CCT and lower corneal hysteresis have been proposed as potential risk and prognostic factors for OAG.^49,50^ These associations have led to the hypothesis that corneal structure and biomechanical properties possibly indicate similar physiological attributes in the ONH. Consistent with this possibility, a small number of *postmortem* non-glaucoma human eyes exhibited similar shear strains in cornea and ONH regions following inflation-induced IOP elevation.^51^ UK Biobank data have also demonstrated relationships among self-reported glaucoma, myopia, and corneal hysteresis.^52^ In contrast, other evidence suggests that the association of CCT with incident OAG may be influenced by the association of CCT with IOP, resulting from collider bias.^53^ Our data found no significant relationships between LC strain and either CCT or corneal hysteresis after accounting for IOP change, and thus do not provide evidence that these corneal measures can serve as direct surrogates for LC strain. A companion report will provide a more detailed analysis of corneal hysteresis and its response to IOP change in relation to LC strain. Longitudinal data from the STRAIN study will elucidate whether CCT and corneal hysteresis are associated with progressive OAG.

### 4.4 LC strain assessment is practical in the clinical setting

Our approach used OCT imaging before and several weeks after temporary washout of existing glaucoma eye drop treatment, initiation of a new eye drop, or SLT to induce IOP change. We then applied DVC to the OCT images to estimate LC strain. This approach can be applied to patients who are already receiving treatment as well as those initiating treatment, using clinically established approaches to alter IOP. For patients initiating new eye drop therapy or undergoing SLT, repeat imaging can be incorporated into routine follow-up without requiring additional visits. For patients already receiving glaucoma treatment, temporary medication washout is an accepted approach in glaucoma clinical trials, with recommendations from the American Academy of Ophthalmology (AAO) and the World Glaucoma Association (WGA).^54,55^ We found that the temporary washout of one or more existing eye drops was acceptable to most patients. As an added measure, home tonometry provided a means for both the patient and research staff to monitor IOP during washout and helped confirm that IOP increased to a sufficient but safe level before repeat imaging. Wider clinical implementation would require automated or streamlined analysis of OCT images, either through software integrated with existing imaging platforms or through an external analysis system. The ability to obtain LC strain measurements during routine glaucoma management could make this approach feasible for longitudinal assessment of ONH biomechanics in clinical practice.

### 4.5 LC strain may be a prognostic factor linked to OAG pathogenesis

Prognostic factors that predict future OAG progression could help guide both the frequency of surveillance and intensity of treatment. Several factors have been proposed as prognostic indicators for OAG, including established clinical factors such as older age and higher mean and variability in IOP, as well as artificial intelligence-based approaches,^56^ polygenic risk scores,^57^ and biomarkers in aqueous humor^58^ or blood.^59^ While these approaches may provide useful information about glaucoma risk or prognosis, LC strain has the potential advantage of directly reflecting the biomechanical environment at the LC, a major site of glaucomatous injury. In particular, LC deformation has been associated with obstruction of axonal transport at this site.^11^ Given this direct mechanistic link, LC strain may be particularly informative if it is found to be associated with future OAG progression. We are currently comparing LC strain responses in STRAIN study participants with their prior visual field and RNFL measurements. Separately, the longitudinal component of the study will determine whether LC strain is associated with visual field and RNFL progression over three years of follow-up.

### 4.6 LC strain may be a modifiable prognostic factor

IOP is the primary modifiable risk factor for OAG, but emerging evidence suggests that modifying the biomechanical response of the LC and surrounding tissues may also have therapeutic potential. Agents that alter LC or scleral biomechanics have been tested,^60,61^ though some interventions have produced adverse effects when scleral strain was reduced.^62^ The biomechanical properties of the ONH are also influenced by biological processes that regulate extracellular matrix composition and remodeling, suggesting another potential avenue for therapeutic intervention. Experimental animal models have suggested beneficial effects of inhibiting transforming growth factor β activity with angiotensin II receptor antagonists (sartans), including reduced RGC injury and loss.^63,64^ Consistent with these findings, some groups of OAG patients taking sartan agents for systemic hypertension have shown lower rates of incident or progressive disease.^65^ If LC strain is found to be a prognostic factor for OAG progression, these findings could help identify the biomechanical states associated with greater or lesser risk of future damage. The ability to measure LC strain in patients with OAG could then provide a means of assessing changes in the biomechanical response in future studies of therapies targeting ONH biomechanics.

### 4.7 Limitations

The study population consisted predominantly of patients with relatively mild OAG who had been treated with topical medications for several years. The study was conducted at a tertiary glaucoma center, and the study population may therefore not be representative of patients seen in other clinical settings or geographic regions. The inclusion criterion requiring at least one eye with a visual field MD > −12 dB resulted in relatively few eyes with severe visual field loss, limiting our ability to evaluate the relationship between LC strain and advanced disease. Further, most IOP changes induced by medication adjustment involved prostaglandin agents, and the effects of these agents on ocular biomechanics are difficult to distinguish from those of the underlying OAG. The present study did not include a non-OAG control group, limiting our ability to determine whether LC strain responses differ between eyes with and without OAG. We are currently estimating LC strain in non-OAG patients following latanoprost-induced IOP reduction, with imaging performed before and after the intervention.

The OCT imaging and DVC methodology also impose limitations on the strain measurements. Because radial OCT scans have lower resolution in the circumferential direction than within the radial-axial imaging plane, strain components involving circumferential deformation have greater uncertainty. Maximum principal and maximum shear strains were therefore evaluated within the radial-axial plane rather than in three dimensions. OCT resolution also does not permit assessment of LC microstructural features, such as individual connective tissue beams, that may contribute to the measured tissue-level strain response. In addition, the present analysis focused on the LC despite the mechanical coupling between the LC and surrounding ocular tissues, particularly the peripapillary sclera (PPS).^66,67^ Future analysis of STRAIN study data will extend strain assessments to the PPS and other posterior ocular tissues.

Finally, the present cross-sectional analysis cannot establish whether LC strain predicts subsequent disease progression. Longitudinal follow-up of the STRAIN study cohort will provide data on the relationship between LC strain and progressive damage among OAG patients. In addition, we will evaluate whether home tonometry measurements before and after IOP alteration are associated with LC strain and OAG damage progression.

### 4.9 Summary

The LC strain response to IOP alteration was estimated in a large cohort of OAG patients following changes in glaucoma medication or SLT. In cross-sectional analysis, axial strain showed a consistent, significant linear relationship with IOP increase and decrease. The quadratic term for the relationship between maximum principal strain and IOP change was significant, and the association showed some asymmetry about zero, suggesting that individual strain measures may reflect different features of the biomechanical response of the LC. Maximum shear strain showed a nonmonotonic pattern across RNFL grades, with significantly lower strain in eyes with abnormal than normal RNFL grading. In combined multivariable models, LC strain remained associated with RNFL thickness, age, axial length, and race after these variables were considered simultaneously with IOP change and other candidate variables. Older age was associated with lower maximum shear and effective strains, longer axial length was associated with lower effective strain, Asian eyes showed greater shear strain than eyes of White participants, and lower RNFL thickness was associated with lower shear strain. We found no significant relationships between LC strain and either CCT or corneal hysteresis. These findings support the feasibility of estimating LC strain in a clinical glaucoma population and provide a baseline for the longitudinal component of the STRAIN study, which will determine whether LC strain is associated with future disease progression.

## Data Availability

The data generated and analyzed during the present study will be made available through Vivli upon publication of the peer-reviewed manuscript.

## Abbreviations

AMD: age-related macular degeneration
CCT: central corneal thickness
DVC: digital volume correlation
GHT: glaucoma hemifield test
ICP: intracranial pressure
IOP: intraocular pressure
LC: lamina cribrosa
LME: linear mixed-effects
MD: mean deviation
OAG: open angle glaucoma
OCT: optical coherence tomography
ONH: optic nerve head
PPS: peripapillary sclera
RGC: retinal ganglion cell
RNFL: retinal nerve fiber layer
SLT: selective laser trabeculoplasty
VFD: visual field defect.

