## Supplement for "Lamina Cribrosa Strain as a Glaucoma Prognostic Factor: The STRAIN Study Methodology and Initial Outcomes"

### Supplemental Table S1: Demographics and clinical measurements

| Variable | | All Data | >20% DVC Corr. | ≤20% DVC Corr. | *p* |
|---|---|---|---|---|---|
| **Patients** | | | | | |
| *n* | | 119 | 105 | 14 | -- |
| Age (years) | | 66.08 ± 11.06 | 66.10 ± 11.24 | 65.93 ± 9.95 | 0.96 |
| Race | Asian | 14 (12%) | 14 (13%) | 0 (0%) | 0.48 |
| | Black or African American | 39 (33%) | 34 (32%) | 5 (36%) | |
| | White | 64 (54%) | 55 (52%) | 9 (64%) | |
| | Other | 2 (2%) | 2 (2%) | 0 (0%) | |
| Sex | Female | 56 (47%) | 53 (50%) | 3 (21%) | **0.04** |
| | Male | 63 (53%) | 52 (50%) | 11 (79%) | |
| Years known OAG | | 8.93 ± 6.80 | 9.13 ± 6.90 | 7.43 ± 5.93 | 0.38 |
| **Eyes** | | | | | |
| *n* | | 205 | 170 | 35 | -- |
| Type | Washout | 153 (75%) | 126 (74%) | 27 (77%) | 0.65 |
| | Wash-in | 39 (19%) | 32 (19%) | 7 (20%) | |
| | SLT | 13 (6%) | 12 (7%) | 1 (3%) | |
| Drug Discontinued (Washout) | Alpha-adrenergic agonist | 6 (4%) | 5 (4%) | 1 (4%) | 0.63 |
| | Beta-blocker | 14 (9%) | 12 (10%) | 2 (7%) | |
| | Carbonic anhydrase inhibitor (CAI) | 2 (1%) | 1 (1%) | 1 (4%) | |
| | Prostaglandin | 95 (62%) | 76 (60%) | 19 (70%) | |
| | Combined beta-blocker and alpha-adrenergic agonist | 2 (1%) | 2 (2%) | 0 (0%) | |
| | Combined CAI and beta-blocker | 25 (16%) | 21 (17%) | 4 (15%) | |
| | Multiple | 9 (6%) | 9 (7%) | 0 (0%) | |
| Drug Started (Wash-in) | Alpha-adrenergic agonist | 6 (15%) | 6 (19%) | 0 (0%) | 0.43 |
| | Beta-blocker | 7 (18%) | 6 (19%) | 1 (14%) | |
| | Carbonic anhydrase inhibitor (CAI) | 2 (5%) | 2 (6%) | 0 (0%) | |
| | Prostaglandin | 14 (36%) | 11 (34%) | 3 (43%) | |
| | Rho kinase inhibitor | 1 (3%) | 1 (3%) | 0 (0%) | |
| | Combined CAI and alpha-adrenergic agonist | 2 (5%) | 1 (3%) | 1 (14%) | |
| | Combined CAI and beta-blocker | 4 (10%) | 2 (6%) | 2 (29%) | |
| | Multiple | 3 (8%) | 3 (9%) | 0 (0%) | |
| Follow-up Time (days) | | 45.66 ± 43.80 | 45.87 ± 43.32 | 44.63 ± 46.69 | 0.79 |
| In-Office iCare | *n* | 160 | 136 | 24 | -- |
| | IOP at Baseline (mm Hg) | 17.60 ± 5.53 | 17.17 ± 4.88 | 19.68 ± 7.74 | 0.06 |
| | Change in IOP (magnitude, mm Hg) | 4.32 ± 3.88 | 4.00 ± 3.58 | 6.12 ± 4.98 | 0.16 |
| Applanation | *n* | 201 | 167 | 34 | -- |
| | IOP at Baseline (mm Hg) | 15.21 ± 4.84 | 14.85 ± 4.70 | 16.97 ± 5.20 | 0.41 |
| | Change in IOP (magnitude, mm Hg) | 4.99 ± 3.58 | 4.93 ± 3.65 | 5.29 ± 3.26 | 0.46 |
| iCare HOME | *n* | 140 | 116 | 24 | -- |
| | IOP at Baseline (mm Hg) | 15.79 ± 4.77 | 15.27 ± 4.12 | 18.38 ± 6.67 | 0.10 |
| | Change in IOP (magnitude, mm Hg) | 3.10 ± 2.75 | 3.05 ± 2.71 | 3.35 ± 2.99 | 0.42 |
| RNFL Average Thickness (μm) | | 69.00 ± 11.99 | 67.92 ± 12.02 | 74.29 ± 10.52 | **0.002** |
| RNFL Average Thickness Grade | Normal | 55 (27%) | 41 (24%) | 14 (40%) | **0.01** |
| | Borderline | 52 (25%) | 39 (23%) | 13 (37%) | |
| | Abnormal | 98 (48%) | 90 (53%) | 8 (23%) | |
| Axial Length (mm) | | 24.81 ± 1.44 | 24.88 ± 1.41 | 24.47 ± 1.55 | 0.92 |
| Central Corneal Thickness (μm) | | 545.55 ± 42.47 | 546.46 ± 42.96 | 541.14 ± 40.29 | 0.70 |
| Corneal Hysteresis (mm Hg) | | 8.79 ± 1.89 | 8.94 ± 1.93 | 8.08 ± 1.55 | **0.04** |
| Mean Deviation (dB) | | -4.71 ± 6.07 | -4.92 ± 6.12 | -3.65 ± 5.81 | 0.13 |
| Glaucoma Hemifield Test | Normal | 55 (27%) | 42 (25%) | 13 (37%) | 0.25 |
| | Borderline | 14 (7%) | 11 (6%) | 3 (9%) | |
| | Abnormal | 136 (66%) | 117 (69%) | 19 (54%) | |
| Optic Disc Area ($mm^2$) | | 1.85 ± 0.48 | 1.89 ± 0.50 | 1.64 ± 0.36 | **0.02** |

*Data expressed as mean ± standard deviation or number (percent). % DVC Corr. = percentage of the LC region with accurate DVC correlation.*

### Supplemental Table S2: Average LC strain errors separated by ΔIOP direction

| | **Washout drop (*n* = 153)** | | | **New drop & SLT (*n* = 52)** | | |
|---|---|---|---|---|---|---|
| | **Mean ± Std** | **Median [IQR]** | ***p*-value*** | **Mean ± Std** | **Median [IQR]** | ***p*-value*** |
| **Average Baseline Error (Displacement: μm; Strain: %)** | | | | | | |
| $U_R$ | 0.004 ± 0.407 | -0.033 [-0.240, 0.215] | -- | -0.043 ± 0.369 | -0.071 [-0.268, 0.153] | -- |
| $U_\Theta$ | -0.035 ± 3.833 | -0.203 [-1.332, 0.993] | -- | 0.083 ± 3.142 | -0.045 [-1.395, 1.442] | -- |
| $U_Z$ | -0.018 ± 0.258 | -0.033 [-0.169, 0.139] | -- | -0.007 ± 0.244 | -0.007 [-0.137, 0.158] | -- |
| $E_{rr}$ | 0.029 ± 0.101 | 0.016 [-0.034, 0.076] | **0.03** | 0.017 ± 0.093 | 0.003 [-0.042, 0.062] | 0.10 |
| $E_{\theta\theta}$ | -0.004 ± 0.144 | -0.014 [-0.087, 0.089] | 0.14 | 0.024 ± 0.148 | 0.003 [-0.063, 0.115] | 0.64 |
| $E_{zz}$ | 0.060 ± 0.158 | 0.035 [-0.014, 0.132] | **<0.001** | 0.039 ± 0.126 | 0.029 [-0.023, 0.086] | **0.002** |
| $E_{rz}$ | 0.016 ± 0.097 | 0.019 [-0.049, 0.078] | 0.99 | 0.008 ± 0.074 | 0.000 [-0.035, 0.055] | 0.72 |
| $E_{r\theta}$ | 0.006 ± 0.098 | 0.012 [-0.050, 0.072] | **0.007** | -0.020 ± 0.095 | -0.013 [-0.083, 0.026] | 0.77 |
| $E_{z\theta}$ | 0.020 ± 0.418 | 0.022 [-0.162, 0.228] | 0.64 | -0.104 ± 0.373 | -0.051 [-0.249, 0.069] | 0.25 |
| **Average Correlation Error (Displacement: μm; Strain: %)** | | | | | | |
| $U_R$ | -0.011 ± 0.073 | -0.001 [-0.045, 0.031] | -- | -0.011 ± 0.105 | -0.001 [-0.038, 0.028] | -- |
| $U_\Theta$ | -0.182 ± 0.351 | -0.189 [-0.375, 0.025] | -- | -0.120 ± 0.356 | -0.089 [-0.325, 0.125] | -- |
| $U_Z$ | 0.620 ± 0.125 | 0.607 [0.543, 0.689] | -- | 0.623 ± 0.118 | 0.607 [0.557, 0.696] | -- |
| $E_{rr}$ | -0.116 ± 0.066 | -0.118 [-0.164, -0.061] | **<0.001** | -0.110 ± 0.067 | -0.115 [-0.175, -0.049] | 0.69 |
| $E_{\theta\theta}$ | 0.248 ± 0.035 | 0.245 [0.227, 0.266] | **0.002** | 0.251 ± 0.033 | 0.257 [0.228, 0.272] | **0.05** |
| $E_{zz}$ | 0.551 ± 0.076 | 0.550 [0.510, 0.596] | **<0.001** | 0.563 ± 0.089 | 0.569 [0.514, 0.636] | 0.28 |
| $E_{rz}$ | 0.002 ± 0.019 | 0.002 [-0.010, 0.015] | 0.74 | 0.001 ± 0.019 | 0.003 [-0.009, 0.009] | 0.81 |
| $E_{r\theta}$ | 0.001 ± 0.015 | -0.001 [-0.007, 0.008] | **0.009** | 0.004 ± 0.015 | 0.003 [-0.007, 0.009] | 0.59 |
| $E_{z\theta}$ | 0.036 ± 0.109 | 0.029 [-0.029, 0.095] | 0.77 | 0.018 ± 0.098 | 0.016 [-0.038, 0.071] | 0.89 |

*Std = standard deviation, IQR = interquartile range. *p-values: linear mixed-effects models vs. average LC strain.*

**Supplemental Table S3: Absolute average LC strain errors**

| | All eyes (*n* = 205) | | |
|---|---|---|---|
| | **Mean ± Std** | **Median [IQR]** | ***p*-value*** |
| **Absolute Average Baseline Error (Displacement: μm; Strain: %)** | | | |
| $U_R$ | 1.573 ± 0.470 | 1.507 [1.250, 1.876] | -- |
| $U_\Theta$ | 3.273 ± 2.825 | 2.348 [1.632, 3.872] | -- |
| $U_Z$ | 0.629 ± 0.254 | 0.585 [0.451, 0.755] | -- |
| $E_{rr}$ | 0.441 ± 0.136 | 0.431 [0.358, 0.527] | 0.74 |
| $E_{\theta\theta}$ | 0.523 ± 0.320 | 0.450 [0.327, 0.579] | **0.001** |
| $E_{zz}$ | 0.236 ± 0.119 | 0.202 [0.163, 0.279] | **<0.001** |
| $E_{rz}$ | 0.270 ± 0.093 | 0.260 [0.208, 0.311] | 0.17 |
| $E_{r\theta}$ | 0.476 ± 0.262 | 0.405 [0.318, 0.529] | **0.007** |
| $E_{z\theta}$ | 0.645 ± 0.462 | 0.512 [0.354, 0.715] | 0.97 |
| **Absolute Average Correlation Error (Displacement: μm; Strain: %)** | | | |
| $U_R$ | 0.646 ± 0.287 | 0.585 [0.452, 0.738] | -- |
| $U_\Theta$ | 1.310 ± 0.602 | 1.200 [0.934, 1.471] | -- |
| $U_Z$ | 0.971 ± 0.168 | 0.943 [0.854, 1.041] | -- |
| $E_{rr}$ | 0.276 ± 0.067 | 0.273 [0.226, 0.325] | **0.001** |
| $E_{\theta\theta}$ | 0.259 ± 0.037 | 0.255 [0.234, 0.279] | **<0.001** |
| $E_{zz}$ | 0.567 ± 0.079 | 0.570 [0.524, 0.615] | **<0.001** |
| $E_{rz}$ | 0.104 ± 0.037 | 0.095 [0.077, 0.122] | **<0.001** |
| $E_{r\theta}$ | 0.097 ± 0.052 | 0.081 [0.062, 0.110] | **<0.001** |
| $E_{z\theta}$ | 0.147 ± 0.082 | 0.130 [0.096, 0.183] | **<0.001** |

*Std = standard deviation, IQR = interquartile range. *p-values: linear mixed-effects models vs. absolute value of average LC strain.*

**Supplemental Table S4: Mean and median pressures and average LC strains for all eyes**

| | Washout drop (*n* = 153) | | | New drop & SLT (*n* = 52) | | |
|---|---|---|---|---|---|---|
| | **Mean ± Std** | **Median [IQR]** | ***p*-value*** | **Mean ± Std** | **Median [IQR]** | ***p*-value*** |
| **Intraocular Pressure (mm Hg)** | | | | | | |
| **Baseline IOP** | 16.5 ± 5.069 | 16.0 [13.0, 19.0] | -- | 20.2 ± 6.2 | 19.5 [16.5, 24.0] | -- |
| **ΔIOP** | 3.2 ± 4.482 | 3.0 [0.0, 6.0] | -- | -4.8 ± 5.3 | -5.0 [-8.0, -3.0] | -- |
| **Average Strain (%)** | | | | | | |
| $E_{rr}$ | 0.171 ± 0.798 | 0.115 [-0.169, 0.325] | **0.02** | -0.086 ± 0.441 | -0.065 [-0.324, 0.169] | 0.14 |
| $E_{\theta\theta}$ | -0.241 ± 1.969 | -0.031 [-0.649, 0.474] | 0.13 | -0.044 ± 1.142 | 0.009 [-0.746, 0.646] | 0.75 |
| $E_{zz}$ | -0.495 ± 1.126 | -0.309 [-0.754, 0.137] | **<0.001** | 0.430 ± 0.954 | 0.276 [-0.176, 0.940] | **0.01** |
| $E_{rz}$ | 0.015 ± 0.488 | 0.011 [-0.172, 0.240] | 0.70 | -0.013 ± 0.408 | -0.035 [-0.289, 0.153] | 0.82 |
| $E_{r\theta}$ | -0.220 ± 1.044 | -0.165 [-0.539, 0.278] | **0.01** | -0.049 ± 0.752 | -0.052 [-0.290, 0.280] | 0.63 |
| $E_{z\theta}$ | 0.054 ± 0.786 | 0.129 [-0.278, 0.434] | 0.40 | 0.033 ± 0.783 | -0.125 [-0.481, 0.470] | 0.76 |
| $E_{max}$ | 1.005 ± 0.984 | 0.849 [0.512, 1.224] | **<0.001** | 1.329 ± 0.868 | 1.174 [0.808, 1.626] | **<0.001** |
| $\Gamma_{max}$ | 1.167 ± 0.746 | 0.964 [0.769, 1.359] | **<0.001** | 1.157 ± 0.548 | 1.048 [0.769, 1.395] | **<0.001** |
| $E_{eff}$ | 4.952 ± 2.916 | 4.341 [3.290, 5.751] | **<0.001** | 4.813 ± 1.784 | 4.328 [3.529, 5.977] | **<0.001** |

*Data for all eyes (n = 205). Std = standard deviation, IQR = interquartile range, ΔIOP = change in IOP. *p-values: linear mixed-effects models vs. zero strain.*

**Supplemental Table S5: Average LC strains by quadrant for all eyes (*n* = 205)**

| (%) | Superior | | Temporal | | Inferior | | Nasal | |
|---|---|---|---|---|---|---|---|---|
| | **Mean ± Std** | ***p*-value*** | **Mean ± Std** | ***p*-value*** | **Mean ± Std** | ***p*-value*** | **Mean ± Std** | ***p*-value*** |
| $E_{rr}$ | 0.436 ± 2.238 | **0.007** | 0.153 ± 0.848 | 0.01 | -0.056 ± 0.849 | 0.48 | 0.067 ± 0.951 | 0.29 |
| $E_{\theta\theta}$ | -0.493 ± 2.649 | **0.009** | -0.116 ± 1.356 | 0.24 | 0.391 ± 2.078 | **0.008** | 0.251 ± 1.889 | 0.07 |
| $E_{zz}$ | -0.391 ± 1.692 | **0.002** | -0.421 ± 1.055 | **<0.001** | -0.552 ± 1.293 | **<0.001** | -0.664 ± 1.362 | **<0.001** |
| $E_{rz}$ | -0.044 ± 1.064 | 0.56 | 0.028 ± 0.639 | 0.53 | 0.011 ± 0.392 | 0.69 | 0.050 ± 0.611 | 0.25 |
| $E_{r\theta}$ | -0.098 ± 0.789 | 0.14 | -0.202 ± 1.394 | 0.06 | -0.083 ± 0.969 | 0.25 | -0.410 ± 1.593 | **0.002** |
| $E_{z\theta}$ | -0.049 ± 1.194 | 0.56 | 0.086 ± 0.979 | 0.21 | 0.174 ± 0.934 | **0.009** | 0.033 ± 0.903 | 0.61 |
| $E_{max}$ | 1.365 ± 2.738 | **<0.001** | 1.064 ± 1.022 | **<0.001** | 0.942 ± 0.932 | **<0.001** | 1.011 ± 1.041 | **<0.001** |
| $\Gamma_{max}$ | 1.288 ± 1.286 | **<0.001** | 1.125 ± 0.657 | **<0.001** | 1.127 ± 0.781 | **<0.001** | 1.192 ± 0.722 | **<0.001** |
| $E_{eff}$ | 5.388 ± 3.972 | **<0.001** | 4.229 ± 2.258 | **<0.001** | 5.510 ± 2.909 | **<0.001** | 4.544 ± 2.307 | **<0.001** |

*To combine and summarize directional strains resulting from both positive and negative pressure changes, strain component values from the smaller group corresponding to negative pressure change (wash-in and SLT, n = 52) were multiplied by -1. Principal strains (maximum principal and maximum shear) and effective strain values were left unchanged. *p-values: linear mixed-effects models vs. zero strain; statistical significance defined as p < 0.0125 after Bonferroni correction for comparison across four quadrants.*

**Supplemental Table S6: Average LC strains by quadrant for eyes with DVC correlation > 20% (*n* = 169)**

| (%) | Superior | | Temporal | | Inferior | | Nasal | |
|---|---|---|---|---|---|---|---|---|
| | **Mean ± Std** | ***p*-value*** | **Mean ± Std** | ***p*-value*** | **Mean ± Std** | ***p*-value*** | **Mean ± Std** | ***p*-value*** |
| $E_{rr}$ | 0.263 ± 0.765 | **<0.001** | 0.080 ± 0.638 | 0.10 | -0.081 ± 0.779 | 0.20 | 0.074 ± 0.853 | 0.26 |
| $E_{\theta\theta}$ | -0.403 ± 1.770 | **0.009** | -0.014 ± 1.189 | 0.88 | 0.327 ± 1.586 | **0.008** | 0.158 ± 1.776 | 0.24 |
| $E_{zz}$ | -0.418 ± 1.198 | **<0.001** | -0.362 ± 0.851 | **<0.001** | -0.432 ± 1.086 | **<0.001** | -0.543 ± 1.213 | **<0.001** |
| $E_{rz}$ | 0.021 ± 0.322 | 0.40 | 0.026 ± 0.552 | 0.54 | 0.016 ± 0.315 | 0.51 | 0.033 ± 0.634 | 0.50 |
| $E_{r\theta}$ | -0.096 ± 0.494 | 0.03 | -0.169 ± 1.093 | 0.05 | -0.112 ± 0.756 | 0.07 | -0.435 ± 1.450 | **<0.001** |
| $E_{z\theta}$ | -0.055 ± 0.824 | 0.38 | 0.068 ± 0.858 | 0.30 | 0.184 ± 0.805 | **0.004** | -0.020 ± 0.783 | 0.79 |
| $E_{max}$ | 1.148 ± 0.855 | **<0.001** | 0.995 ± 0.724 | **<0.001** | 0.893 ± 0.771 | **<0.001** | 1.038 ± 0.952 | **<0.001** |
| $\Gamma_{max}$ | 1.158 ± 0.700 | **<0.001** | 1.045 ± 0.554 | **<0.001** | 1.048 ± 0.697 | **<0.001** | 1.153 ± 0.685 | **<0.001** |
| $E_{eff}$ | 4.887 ± 2.259 | **<0.001** | 4.007 ± 1.905 | **<0.001** | 5.307 ± 2.686 | **<0.001** | 4.450 ± 2.242 | **<0.001** |

*To combine and summarize directional strains resulting from both positive and negative pressure changes, strain component values from the smaller group corresponding to negative pressure change (wash-in and SLT, n = 44) were multiplied by -1. Principal strains (maximum principal and maximum shear) and effective strain values were left unchanged. *p-values: linear mixed-effects models vs. zero strain; statistical significance defined as p < 0.0125 after Bonferroni correction for comparison across four quadrants.*

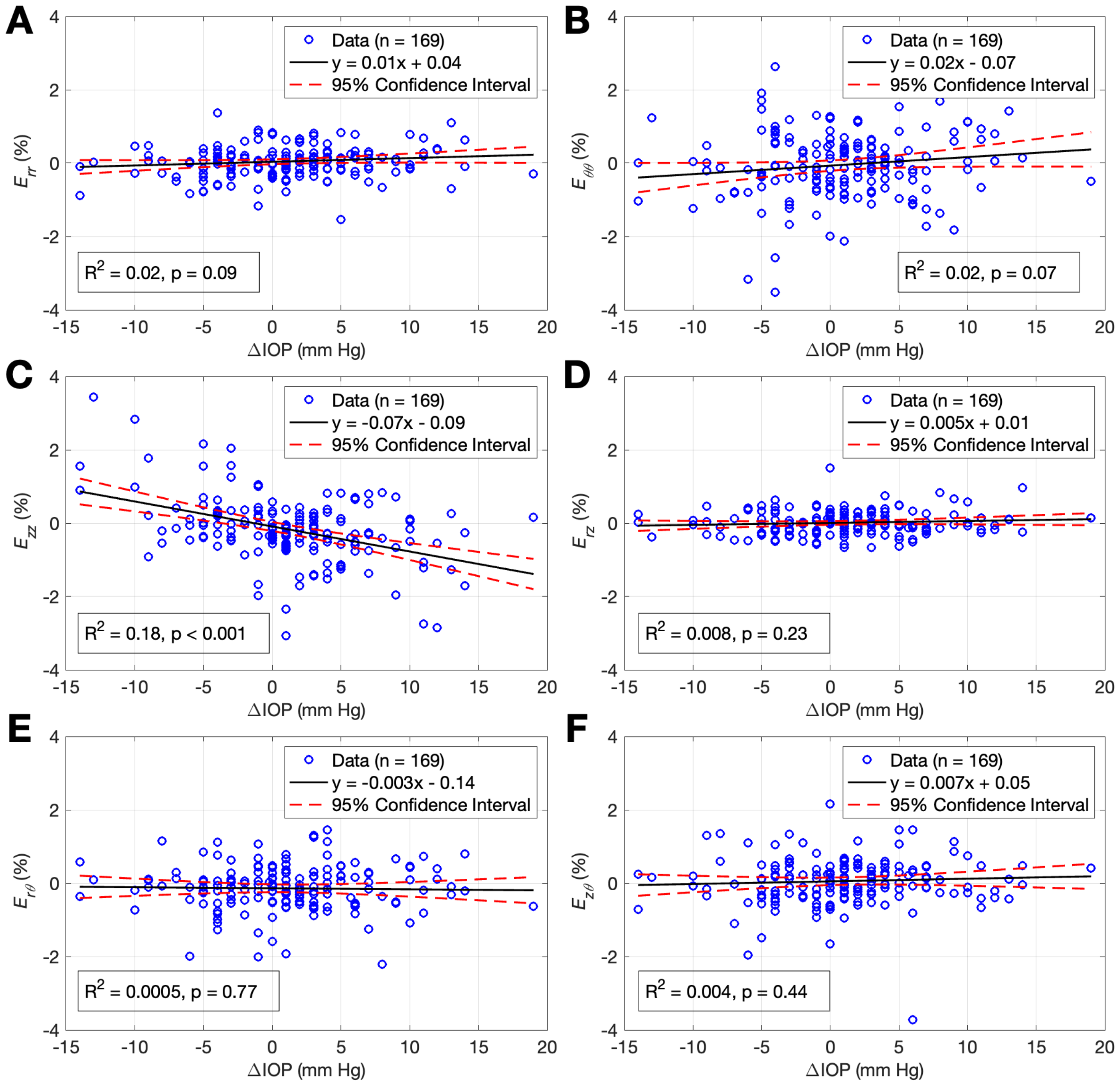


**Supplemental Figure S1: LC strain components versus IOP change.** (A) Radial strain ($E_{rr}$), (B) circumferential strain ($E_{\theta\theta}$), (C) axial strain ($E_{zz}$), (D) radial-axial shear strain ($E_{rz}$), (E) radial-circumferential shear strain ($E_{r\theta}$), and (F) axial-circumferential shear strain ($E_{z\theta}$) versus IOP change. Axial strain showed a significant linear relationship with IOP change, with similar strain magnitudes observed for IOP increases and decreases of similar magnitude. Radial and circumferential strain also showed linear relationships with IOP change, though these relationships did not reach statistical significance. Shear strain components were not associated with IOP change.

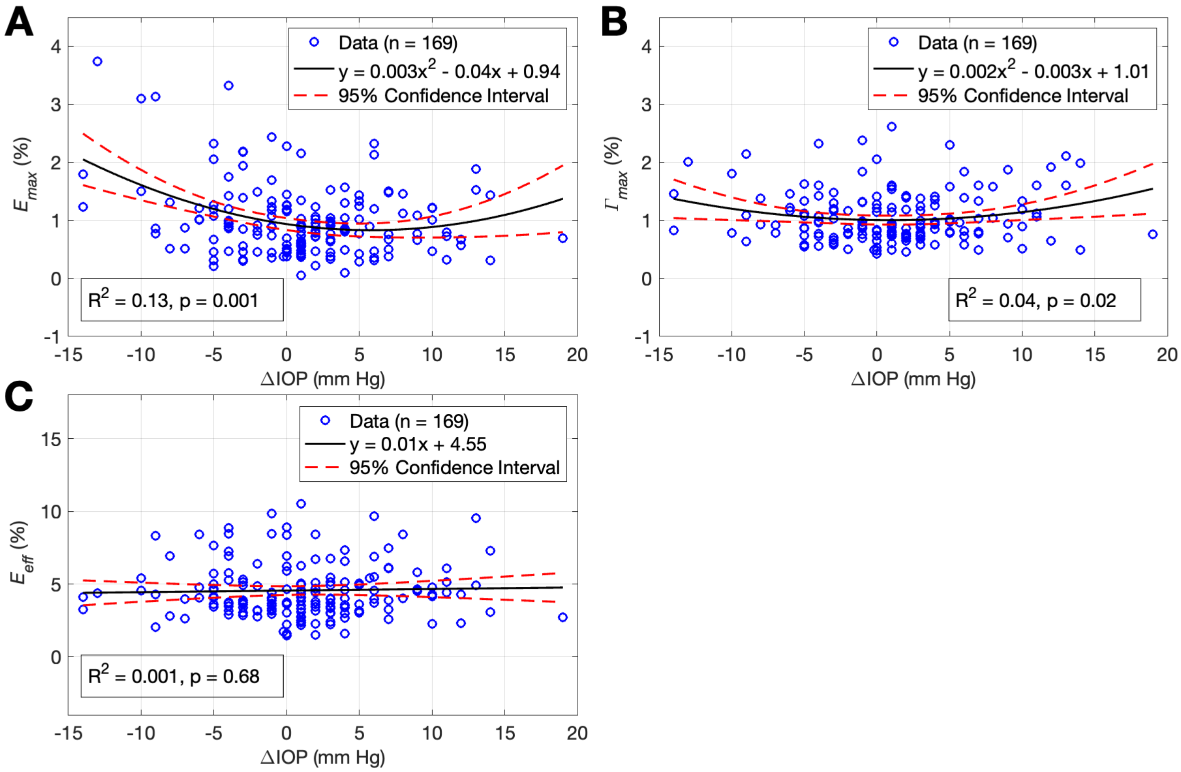


**Supplemental Figure S2: LC strain versus IOP change.** (A) Maximum principal strain in the axial-radial plane ($E_{max}$), (B) maximum shear strain in the axial-radial plane ($\Gamma_{max}$), and (C) effective strain ($E_{eff}$) versus IOP change. Maximum principal strain showed a significant quadratic relationship with IOP change that was not symmetric around zero. Maximum shear strain and effective strain were not associated with IOP change.
